# INTERVENTIONS TO IMPROVE PATIENT SAFETY DURING THE COVID-19 PANDEMIC: A SYSTEMATIC REVIEW

**DOI:** 10.1101/2024.06.10.24308558

**Authors:** AW Wu, K Trigg, A Zhang, GC Alexander, ER Haut, C Rock, KM McDonald, WV Padula, S Fisseha, R Duncan, J Black, DE Newman-Toker, I Papieva, N Dhingra, R Wilson

**Author notes:** **Ethical Approval** Not Applicable.

## Abstract

**Objective:** To summarize the literature on health care interventions to reduce harm to patients caused by the COVID-19 pandemic across six domains: medication errors, diagnostic errors, surgical errors, health care-associated infections, pressure injuries, and falls.

**Methods:** We performed a mixed methods systematic review, with the intention to present results narratively. We combined parallel searches and experiential evidence across each domain of interest. We included studies published between 11 March 2020 and 28 August 2023 that reported an intervention in response to an identified patient safety issue. We identified 13,019 unique articles across the six domains. Of these, 590 full texts were assessed for eligibility. Seven were included for the medication safety domain; seven for diagnostic safety; 32 for surgical safety; 11 for health care-associated infections; six for the pressure injuries; and two for falls (Annex C). Overall, a total of 61 unique articles were included – four articles were represented across more than one domain.

**Findings:** There were few rigorous evaluations of specific interventions to reduce patient harm caused by the pandemic. Adjustments in treatments, triage, and procedures, and use of risk stratification tools reduced delays and permitted more elective surgery and diagnostic testing to proceed, improvements in medication safety practices, and prevention of health care-associated infections. Publications emphasized the importance of implementing existing practices and following the latest guidelines to prevent health care-associated infections, medication errors, pressure injuries and falls.

**Conclusion:** There is little research on interventions to reduce patient harm caused in health care settings during the COVID-19 pandemic. Interventions focused on preventing nosocomial transmission of COVID-19, and on permitting access to urgent surgical and diagnostic needs. A few studies tested strategies to reduce new risks imposed by the pandemic for medication errors, health care-associated infections, pressure injuries, and falls. They also urged extra efforts to implement existing practices and following the latest guidelines already known to be effective. Development of high-reliability health systems and health care organizations to protect patients and health workers from harm, will be essential to mitigating the impact of future pandemics within the objectives of the Global Patient Safety Action Plan 2021-2030.

## INTRODUCTION

The COVID-19 pandemic presented an unprecedented challenge for countries around the world, testing their health systems’ resilience to navigate through unknown, adapt to the constantly evolving situation and respond in a coordinated and cooperative manner. Its toll on the global population was staggering, with 18 million estimated deaths at the beginning of 2023^1^. Funding was inadequate, particularly for low and middle-income countries. Supplies and distribution of key commodities including protective equipment, diagnostic tests, devices, medications and vaccines were insufficient and inequitably distributed. Furthermore, there was a lack of timely, accurate and systematic data on infections, health consequences and responses^2,3^.

Patient safety is a crucial issue for health systems around the world and essential for ensuring high-quality health care for making progress towards universal health coverage and achieving the United Nations Sustainable Development Goals ^4,5^ Unsafe health care can be a tragedy for individual patients and their families, and a major impediment to success for communities and national economies^6^

Prior to the COVID-19 pandemic, there were demonstrated challenges with patient safety in countries^2^. In low- and middle-income countries, unsafe and poor-quality care were greater causes of preventable death than problems with access to care^7^ The pandemic spotlighted gaps in patient and health worker safety across several domains, with increases in specific types of harm^8^. There were increases in health care-associated infections from COVID-19 and other agents due to non-compliance with existing precautions, increases in medication errors and unsafe medication practices due to inadequate communication and drug-to-drug interactions, harm due to delays in surgical care and diagnostic testing, and increases in pressure injuries and falls due decreases in patient monitoring. These specific safety domains are the focus of this review.

## METHODS

We conducted a narrative systematic review using mixed methods, employing parallel searches of PubMed, Embase, CINAHL, and APA PsycINFO and incorporation of experiential evidence. We addressed the selected high-risk areas identified in the rapid review conducted by WHO in 2022 ^8^. Studies were included and excluded based on the general criteria identified in our PICOTS framework (Table 2).

### Search Strategy

Searches were initially developed and tested in PubMed, then adapted to the other databases (Appendix 1-Appendix 6). Terms were developed for the specific domains in consultation with our experts. Searches for all domains also included terms relevant to patient safety, and terms related to patient harm. All searches included a string designed to exclude non-interventional studies such as case reports, editorials, commentary, and perspective pieces. Meeting abstracts were not included due to the sparsity of data they present. Systematic reviews were not included, but those that were relevant were tagged and their references were screened for applicability. Searches were not restricted by language but were restricted by date: 11 March 2020 to 28 August 2023.

Searches for medication safety and surgical safety returned a large number of potential titles and abstracts. These two searches were further restricted by a set of terms specific to COVID-19 (Annexes A1 and A3). The search for surgical safety search (Annex A3) was amended to better capture studies reporting on surgical delays which can be considered a patient safety issue. Interventions were categorized by system components/elements from the adapted WHO health system framework. These included interventions related to: health services; the health and safety of health workers; patients, families and communities including inequities; leadership, governance and financing; communication and management of health information; and development and supply chain of medical products, vaccines and technologies.

### Screening and Abstraction

Titles and abstracts were deduplicated and uploaded to DistillerSR (DistillerSR. Version 2.35. DistillerSR Inc.; 2021) for screening. Each title/abstract was independently screened by two individuals. Agreement between the two screeners was required for a title/abstract to be either excluded or included (moved to the next level of screening). Disagreements that could not be resolved between the two screeners were referred to a third screener for adjudication. The same screening protocol was applied to the full texts. For all included studies and grey literature, domain leads extracted country, practice and participant characteristics, and intervention data (Annex B).

### Extraction

Data from the included studies were compiled into evidence tables (Annex D) and a narrative synthesis was performed of the findings extracted from the primary studies.^10^

#### Analysis

After data was extracted, two team members independently identified themes within each study. We then reviewed these initial themes and combined them into common themes. Themes were grouped by reviewing short synopses of studies to determine appropriate categories. Results are synthesized narratively. Studies are listed under thematic areas irrespective of independent determination, where independent reviews were included and did not require double adjudication. We recorded the overall number of studies and studies and intervention themes associated with each safety domain.

### Assessment of Risk of Bias

We assessed individual study risk of bias using tools appropriate for study design. We used the Risk of Bias 2 (RoB2)^10^ tool for randomized studies, the Newcastle-Ottawa Scale for non-randomized cohort studies and case-control studies^11^. No alterations to the tools were considered and the established scoring algorithms were used. We did not conduct evidence grading as this is not feasible in a narrative review.

This narrative review was assessed using the Scale for the Assessment of Narrative Review Articles (SANRA)^12^ (Annex F).

## RESULTS

There were 13,019 unique articles identified across the six safety domains. Of these, 590 were assessed for eligibility. Overall, a total of 61articles were included. However, several studies spanned across multiple domains. Studies that crossed multiple domains were included in each domain and were independently reviewed. Seven were included in the medication safety domain; seven in the diagnostic safety domain; 32 in the surgical safety domain; 11 in the health care-associated infections domain; six in the pressure injury domain; and two in the falls domain (Annex C).

### Safety Domains

#### Medication Errors

Studies on medication safety covered five themes across seven studies: adapted safety guidelines (1), modified procedures (1), team communication (5), information technology (3), and education (1). Facilitating team communication was common across these studies, with different interventions such as development of a modified pathways to determine the safety and feasibility ^13,14–16^ Information technology was described in three studies to prevent prescribing errors^13^.

There were also studies which implemented software to reduce drug-drug interactions^14^ and risk scoring to simulate interventions for repurposing drugs^15^ (all of which were associated with an increased risk of adverse drug events). One study proposed the use of education to improve the awareness and attitudes towards adverse drug reaction reporting^16^.

All seven studies were assessed as “fair” using the Newcastle-Ottawa scale for cohort studies (see Annex E, Table 3). Reasons for downgrading the studies: 1 study selectively recruited participants; 5 of the studies did not include a control group; 1 study did not have adequate follow up. Annex E, Table 3 includes details of the individual study assessment.

#### Diagnostic Errors

Studies on diagnostic safety covered five themes across the seven studies: triaging (1), adapted safety guidelines (2), modified procedures (4), information technology (5) and telemedicine (2). One study examining implications of the delays in cancer referral on outcomes found that test triaging has short-term benefits such as streamlining access^17^. Two studies discussed interventions devised to increase patient safety during the diagnostic process via early testing and a risk-stratified approach^18,20^. Additional interventions included variations of telehealth including teleconsultation^19^ and homecare instead of outpatient care^20^. One study used a thromboprophylaxis algorithm to reduce the risk of thrombotic events in patients with COVID-19^21^.

All seven studies were assessed as “fair” using the Newcastle-Ottawa scale for cohort studies (see Annex E, Table 3). Reasons for downgrading the studies: 5 of the studies selectively recruited participants; none of the studies identified a control group; follow up was not adequately defined in any of the studies. Annex table 3 includes details of the individual study assessment.

#### Surgical Errors

Studies on surgical safety covered seven themes across the 32 included studies: triaging (14), adapted safety guidelines (17), modified procedures (25), team communication (3), information technology (9), education (4), and telemedicine (6). All of the themes identified were included in multiple studies. For example, 14 studies employed triaging and modified patient pathways which included creation of clean surgical sites^22^, outpatient rather than inpatient care^23^, novel approaches to ambulatory surgeries^24^, COVID-19-minimal pathways^25–28^, and operating room modifications^29–31^. Studies also examined redesign/reorganization^32–35^, prioritization protocols^36^ and pivot plans for resumption of services^37^. Examples of studies which utilized risk stratification^38,39^ and information technology strategies^7,28,42^ includes testing protocols based on guidelines and input from interdepartmental team members^40,41^. One study created a protocol for international medical missions to mitigate exposure and transmission risk and implemented telemedicine, education, and modified procedures in alternative follow-up protocols^42^. Studies also included the use of alternative and modified institutional protocols and safety measures^43^. Several studies described novel tools and techniques such as devices to continue performing percutaneous tracheostomies^44,45^, low-cost filtration devices^46^, and conservative management of appendicitis^45,47^.

Thirty-one studies were assessed using the Newcastle-Ottawa scale for cohort studies, four of these studies were assessed as “good” and one was assessed using the Newcastle-Ottawa scale for case-control studies, and was assessed as “fair.” (see Annex E, Table 4). Reasons for downgrading the remaining cohort studies: 3 studies did not describe the exposed cohort, and 9 studies had a somewhat representative cohort of participants or selectively recruited participants; 24 studies did not describe what elements were controlled for in the analyses; 6 studies did not define how outcomes were assessed, and 10 did not describe the number of participants who completed the study or had an unacceptable loss to follow up. The case control study was downgraded for using record linkage and hospital controls. Annex E, Table 4 includes details of the individual study assessment.

#### Heath Care-Associated Infections (HAI)

Studies on health care-associated infections covered seven themes across the 11 studies: triaging (3), adapted safety guidelines (2), modified procedures (9), team communication (3), information technology (3), education (2), and telemedicine (2). In the largest thematic area, modified procedures studies included the use of short sleeve gowns^48,49^, early discharge^49^, use of respiratory drive throughs^50^ and enhanced multimodal infectious prevention and control strategies to reduce transmission^51^. Studies also examined innovations such as adapted pathways^43,50^, triaging, incorporating patient liaisons, and telemedicine^50^. Studies described nurse-led quality initiatives in the form of nurse-led toolkits^52,53^, a multidisciplinary stakeholder team^53^, and a training model^54^. One study also described using respiratory hygiene procedures like those used during influenza epidemics to reduce COVID-19 transmission^49^. Bundling technology together with education was evaluated to reduce central line-associated blood stream infections and catheter-associated urinary tract infections^55^. There were also several technological innovations presented such as the use of visualization techniques to view rates of transmission^56^.

Eight of the 11 included studies were assessed using the Newcastle-Ottawa scale for cohort studies, the 3 unassessed studies were descriptions of quality improvement projects studies (see Annex E, Table 3). Of the assessed studies, three were scored as “good” and the remainder, “fair.” Reasons for downgrading the assessed studies: one study selectively recruited participants, and three studies did not describe the unexposed cohort; 6 studies did not describe what elements were controlled for in the analyses; 3 studies gathered data through self-report. Annex E, Table 3 includes details of the individual study assessment.

#### Prevention of Pressure Injuries

Prevention of pressure injuries covered six themes across six studies: triaging (4), adapted safety guidelines (1), modified procedures (5), team communication (3), information technology (1), and education (1). Multidisciplinary and interprofessional teams were developed in one study with the goal of reducing workloads and standardizing processes^57^, while another study described establishment of a dedicated team to handle tracheostomies^58^. Two studies used novel pathway management by reorganizing the ICU^59^ and developing a prone positioning protocol and strategy^60^. One study found that reallocation of healthcare workers to perform prone positioning outside of their typical scope of work was feasible and safe^61^.

Four of the six included studies were assessed using the Newcastle-Ottawa scale for cohort studies and all studies were assessed as “fair”; one study was assessed using Cochrane’s Risk of Bias 2, and had a low risk of bias, the one unassessed study was a description of quality improvement (see Annex E, Table 5). The cohort studies were downgraded for the following reasons: 2 studies did not describe the unexposed cohort; there was no description of elements controlled for in analyses; 2 studies gathered data through self-report. Annex E, Table 5 includes details of the individual study assessment.

#### Prevention of Falls

Two studies across three themes covered falls prevention: triage (2), modified procedures (2), and team communication (1). Both studies on falls prevention discussed different triaging and modified procedures such as ICU reorganization^59^. Another study discussed the use of team communication and collaboration to identify patients at increased risk^62^.

Two studies were assessed using the Newcastle-Ottawa scale for cohort studies and all studies were assessed as “fair” (see Annex E, Table 3). Reasons for downgrading include: studies did not describe what elements were controlled for in the analyses; 1 study gathered data through self-report. Annex E, Table 3 includes details of the individual study assessment.

### Interventions

Across the six safety domains, we identified seven intervention themes: triaging (including novel triage, pathway triage, and care pathway innovations); adapted safety guidelines (including safety protocols and guidelines; risk scoring and prediction; PPE, and hygiene measures); modified procedures (including adapted isolation, enhanced ventilation); team communication (text messaging, committees); information technology (including simulations, and software); education; and telemedicine. The themes that appeared in each domain are shown in Table 1. There were thematic similarities across the safety domains.

#### Triaging and Care Pathways

Twenty-one studies across five domains assessed novel pathways or triaging of care. Of these, care pathways were employed to improve surgical safety and pressure injury and falls. The use of remodeled surgical^25,27,36,58^ pathways, streamlined pathways^26^ and mitigating plans^29,30,59^, along with reorganization of units^25,59^ were suggested to improve patient acuity and reduce staffing burden. Hygiene procedures were modified^49^ and in-home testing for colonoscopy^17^ was incorporated to improve diagnostic safety. An intercom system was used to improve communication about activation of patient call lights^62^ and prevent pressure injury and falls. Telemedicine^63^ was suggested as an alternative mechanism for following surgical patients. There was no overlap in the types of intervention used to increase patient safety within the triaging domain, except for in two studies^25,26^ that described comparable streamlined surgical pathways.

#### Adapted Safety Guidelines

There were 21 studies that used adapted safety guidelines to prevent harm in five safety domains. For HAIs, two studies utilized a nurse-led toolkit^52,53^ to prevent infections. A unique, pharmacologic approach to stress testing ^18^ was used to improve diagnostic safety. Novel guidelines were implemented across four studies spanning pressure injuries and falls^58^ and surgical safety^30,36,38^, and using a pivot plan^37^ to allow resumption of elective surgeries. The pivot plan was developed to help find safe pathways for elective surgeries during a pandemic, and consisted of guidance to support a resumption of services across procedural areas. One surgical study evaluated making a change in surgical selection criteria^34^.

#### Modified procedures

There were 42 studies that assessed the use of various modified procedures across all safety domains. Many of these studies also described other intervention themes, such as the use of modified safety checklists^35^, care pathways^17,25,42^, environmental modification^64^, and changes in procedures and hygiene practices^49,51^. One study evaluated management of pharmaceutical interventions and added pharmacists to the team to reduce medication errors^65^. Another study revised follow up procedures of patients in conjunction with a new protocol for nuclear imaging^18^.

#### Team Communications

There were 14 studies that evaluated team communications to improve 5 safety domains. Within HAIs, the nurse-led toolkit^53^ included adapted communication guidelines to prevent bloodstream infections^55^. For medication safety three studies assessed the addition of pharmacists^13,65,66^ and improved communication among teams^66^ to reduce errors. One novel study used WhatsApp as a digital platform to improve communication^13^. For pressure injury and falls there was research on creating new teams to improve communication^57,62^. Medication safety interventions most often emphasized teamwork and communication, primarily using pharmacy teams.

#### Information Technology

Twenty studies used information technology such as software, risk scoring or prediction and simulation strategies to improve patient safety in five domains. In one study, natural language processing was used to identify and categorize diagnostic errors by reviewing all patient safety reports mentioning COVID-19 and identifying additional safety reports on errors or delays^67^. Within medication safety, one study noted above used WhatsApp to share prescribing error scenarios among community pharmacists^13^ while another evaluated the use of data-based software to check for Drug-Drug Interactions^14^. The three surgical safety studies creating modified risk-scoring^38,68^ along with defined selection criteria^34^ to determine candidacy for surgery.

#### Education

Seven studies used educational interventions in four safety domains. Education on hygiene and distancing measures^49^, staff education regarding new or adapted protocols^27^, and team development^57,60,62^ were most common.

#### Telemedicine

The use of telemedicine was not as prevalent as expected in this literature, with only 10 studies explicitly utilizing telemedicine to improve safety in three domains. These primarily incorporated modified procedures^19,20,49^, novel pathways^24,42,50^, and patient management^27,63^ following adaptations to procedures.

**Table 1.**
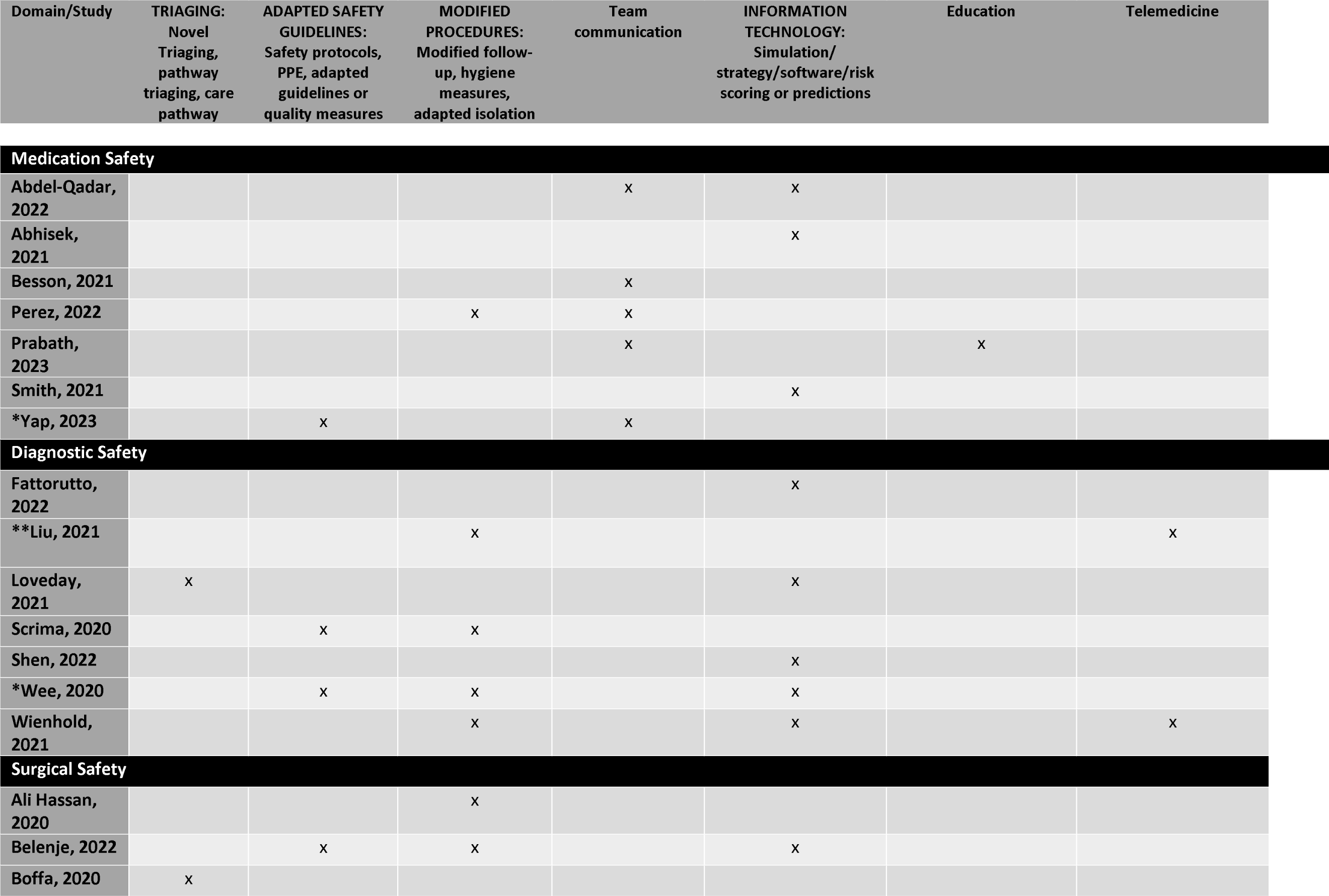
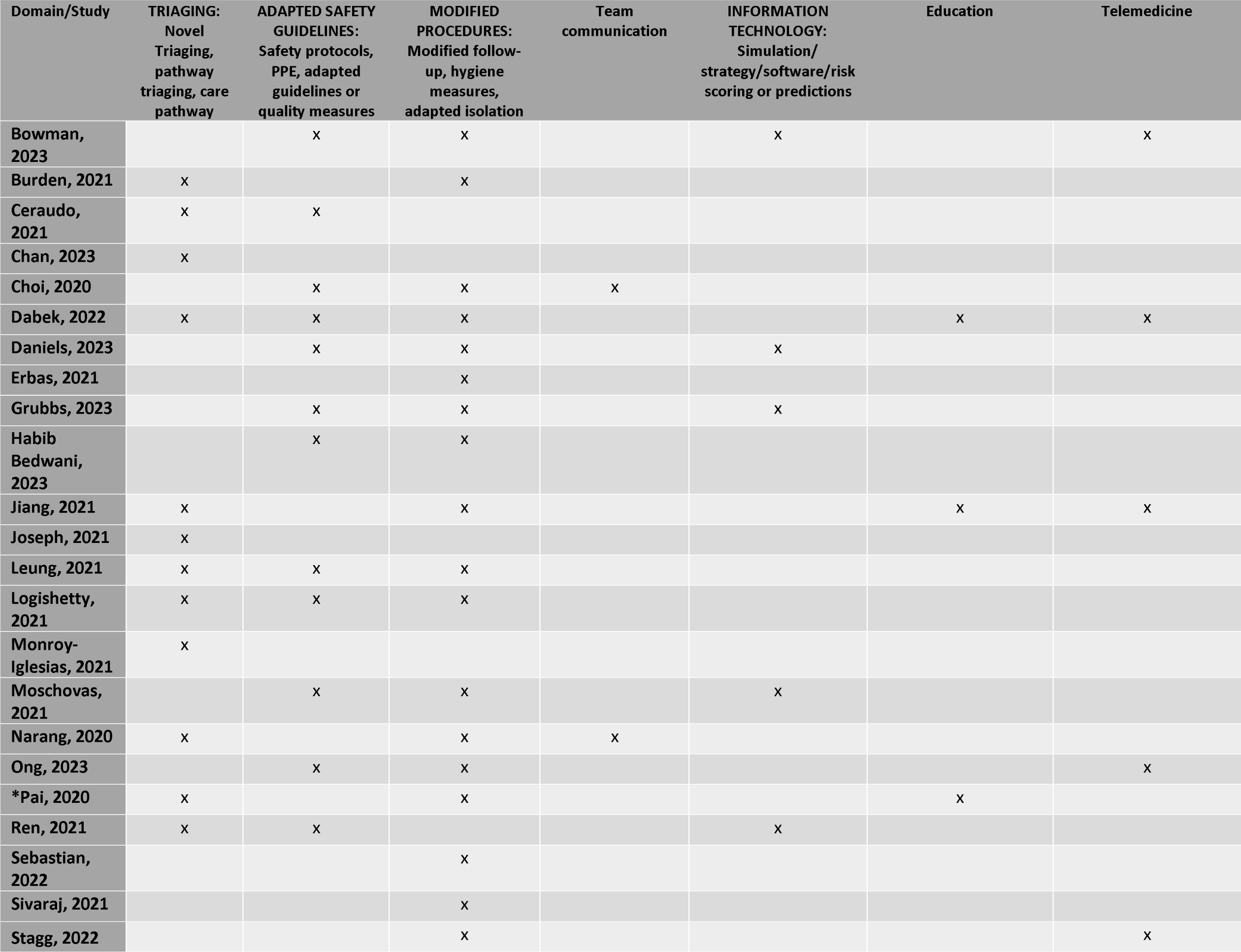
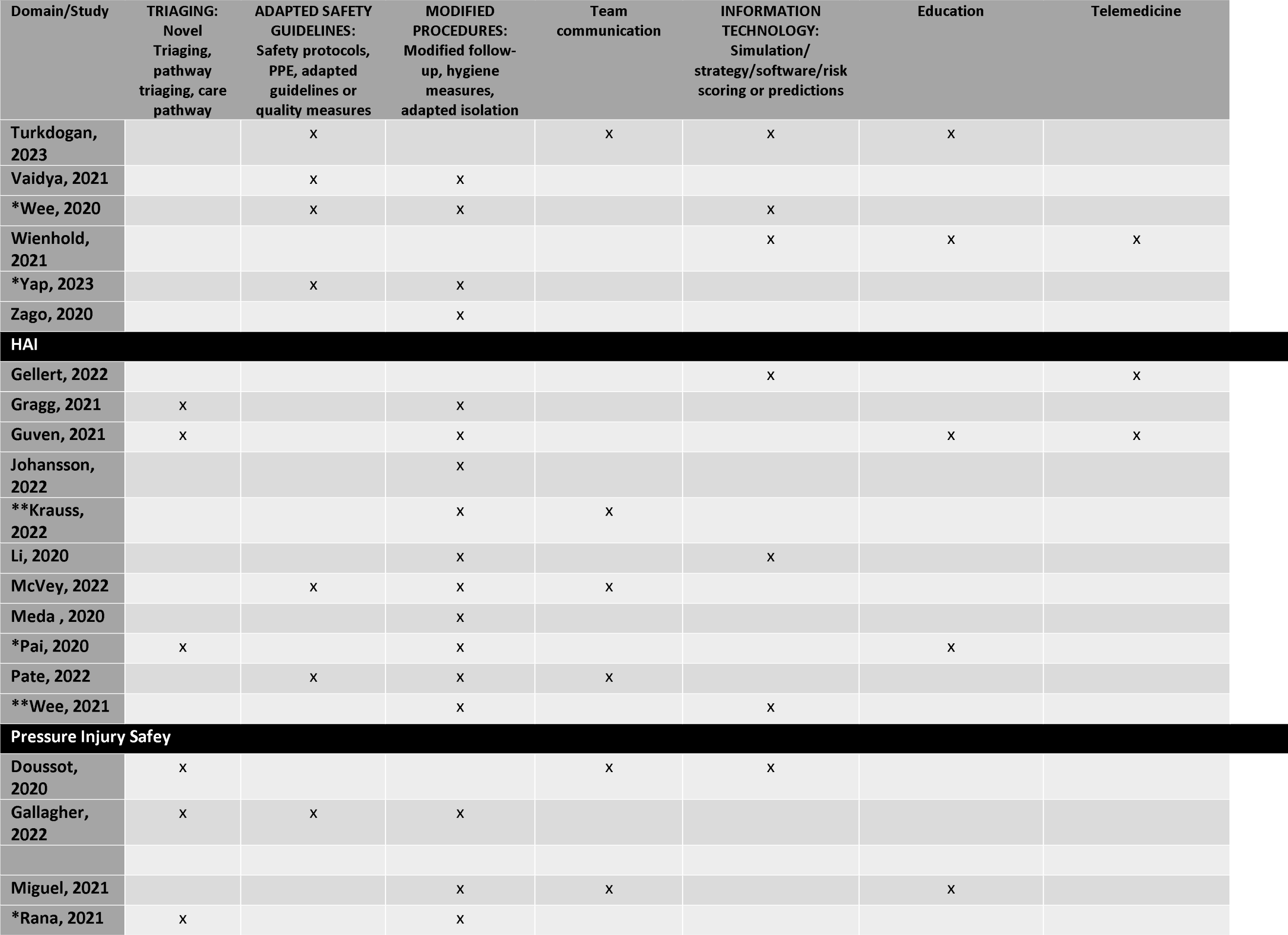
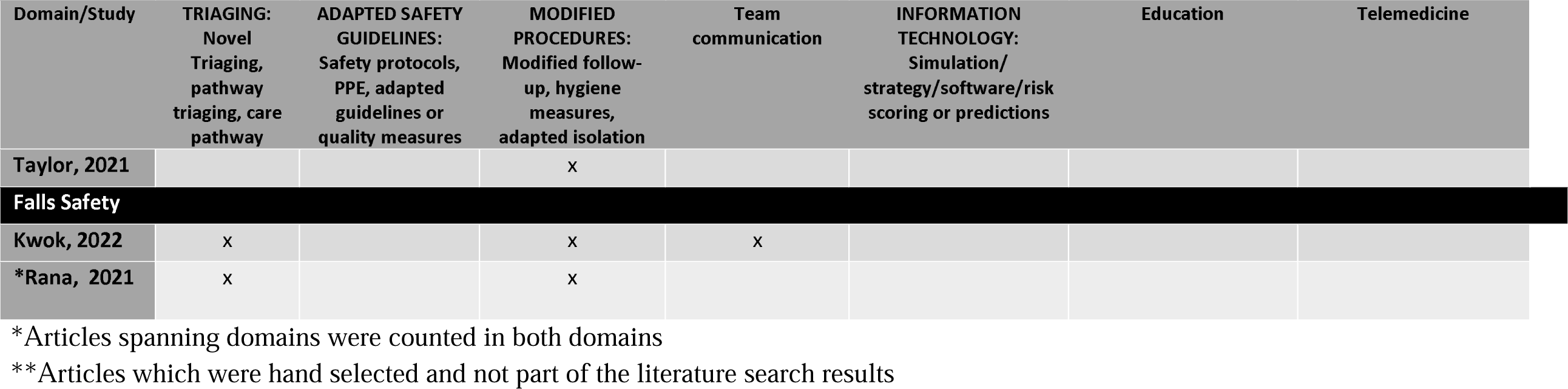
Intervention themes appearing within each domain (n)

## DISCUSSION

Our findings provide a systematic summary of the literature on interventions to prevent or mitigate patient safety challenges caused by the COVID-19 pandemic. In general, relatively few studies assessed or described these interventions.

The largest number of studies addressed surgical safety. Beginning early in the pandemic, efforts to reduce contagion led to drastic restrictions on surgical capacity. Performing surgery safely on patients with COVID-19 was a challenge for providers and health systems, and changes were made to treatment approaches in surgical diseases to reduce transmission risk. However, the secondary impact of the pandemic on uninfected surgical patients was greater and drove changes in surgical care. Accurate triage procedures were critical to identifying patients most urgently in need of surgery, and multiple interventions relied on systematic approaches to prioritize individuals.

Few studies addressed interventions to improve diagnostic safety. Studies on diagnosis focused on tools to increase the safety of individual patients and providers rather than to reduce diagnostic errors. A few attempted to prevent delays in time-sensitive diagnostic evaluation. Further research on the short-term and long-term outcomes in patients undergoing surgical care and diagnostic evaluation needs to be conducted to ensure these interventions improve safety.

There were few studies of interventions to improve medication safety. These focused primarily on the reduction of medication errors through innovations in communication and information technology. Medication errors have been identified as perhaps the most common source of adverse events for patients worldwide^69,70^. Medication errors have received extensive study and elicited a series of recommendations including those from the Third WHO Global Patient Safety Challenge: *Medication Without Harm* and the Global Patient Safety Action Plan 2021-2030^71–75^. Thus, the lack of new interventions may not be surprising, with authors and experts recommending instead an emphasis on complying with existing evidence-based practices.

The same may be said for interventions to prevent and reduce health care-associated infections during the pandemic. Reducing the transmission of SARS-CoV-2 itself was a primary goal of any health system. Prevention of other prevalent health care-associated infections was addressed with innovations such as toolkits as well as enhanced personal protective equipment, and hygiene measures. Researchers and experts emphasized compliance with existing guidelines and best practices dating back to the WHO Clean Care is Safer Care campaign,.

The small number of studies to address pressure injuries and patient falls focused on active interventions and physical measures to decrease incidence of these types of harm. In addition, authors and experts recommended compliance with existing best practices for the prevention of pressure injuries^80^ and hospital falls^81,82^. The literature on falls emphasizes that patients and families should be involved in planning and implementing the fall prevention process^83,84^.

Many of the recommendations to protect patients and health workers that pre-dated the COVID-19 pandemic are incorporated in the Global Patient Safety Action Plan 2021-2023^72^. In addition to specific actions related to practice improvements and strengthening policy environment, the plan emphasizes the importance of building high-reliability health systems and health care organizations. One of the key objectives is to partner with patients and families, and engage and empower them to be proactive partners in their own care. This includes working with them to ensure shared decision-making at the point of care, co-developing guidelines and procedures as well as contributing to policy development and health system design and governance to make health care safer.

### Limitations

This study had some limitations. Based on the narrow time period of this study and lag in the publication of research, we are likely to have missed results that are still forthcoming. We did not expect to find many new interventions to improve patient safety across the researched safety domains. However, the presence of common themes across interventions indicates that health workers did make use of existing evidence and experience to reduce errors and improve safety.

### Summary and Implications

The findings of this review highlight avenues and methods that can be followed and applied in future outbreaks and pandemics. There was an overriding imperative to prevent nosocomial transmission of COVID-19, and several of the interventions focused on the protection of patients, family members and health care workers. Beyond this, there was a compelling need to treat patients in need of urgent and emergent surgical care, and to provide timely diagnostic evaluation for people with other medical problems. This was necessary to prevent a secondary epidemic of morbidity due to preventable and treatable conventional conditions. A second group of interventions focused on providing access to these services. Several new interventions focused on patients secondarily exposed to increased risk from common problems including medication errors, health care-associated infections, pressure injuries, and falls. However, the authors of these papers primarily urged extra efforts to implement existing practices and follow latest guidelines already shown to be effective.

Overall, there has been relatively little research on interventions to reduce patient harm due to health care during the COVID-19 pandemic. There is a need for additional evidence on effective interventions that can be implemented by health workers and patients, and health systems in general at national, sub-national and institutional levels. Some of the necessary competencies can be adopted and adapted from existing knowledge and experiential evidence. The Global Patient Safety Action Plan 2021-2030^72^, provides a foundational framework of strategies and actions for all stakeholders that will help to mitigate the impact of future outbreaks and pandemics on patient safety. A goal will be the development and adoption of sustainable improvements within high-reliability health systems and health care organizations that can protect patients and health workers from harm^85^. Partnering with patients and families will encourage their engagement and empowerment in reducing harm.

## Supporting information

Annex E

Annex D

Annex B

Annex C

Annex A

Annex F

## Data Availability

All data produced in the present work are contained in the manuscript

## Acknowledgements

The authors would like to acknowledge the contributions of Lindsay Bow, Erin Barker, Amber Thomas, Dillu Bajgai, the Armstrong Institute Center for Diagnostic Excellence for supporting the effort on this study, and the World Health Organization’s Patient Safety Flagship Unit for supporting this systematic review.

---

**Table 2.** Inclusion and exclusion criteria based on population, interventions, comparators, outcomes, timing, and setting.□.

| PICOTS | Inclusion criteria | Exclusion criteria |
| --- | --- | --- |
| Populations | Global—populations (any) impacted by a pandemic | Studies with a population less than 5 participants |
| Intervention | Safety/quality measures—for patients<br>Safety/quality measures—for health and care workers and associated services<br>Prevention measures<br>Treatments<br>Vaccines<br>Care transformation | Interventions that do not impact or increase patient safety |
| Comparators | NA | NA |
| Outcomes | General:<br>Mortality<br>Long-term morbidity<br>Treatment delays<br>Long-term disruptions in preventive care<br>Health and safety of health care workers<br>Mental health<br>Inequity<br>Domain specific:<br>Medication safety<br>Diagnostic safety<br>Surgical safety<br>Health care associated infections<br>In-patient health concerns (pressure injury and falls) | Does not discuss or describe the listed outcomes of interest<br>Studies on patient management |
| Timing | Studies completed during the COVID-19 pandemic* | Studies completed prior to the COVID-19 |
|  |  | pandemic ☐ |
| Setting ☐ | All care settings (medication, diagnostic, surgical, health care associated infection) ☐<br>Inpatient setting (pressure injury and falls) ☐ | None ☐ |

PICOTS = populations, interventions, comparators, outcomes, timing, setting ☐
\*WHO declared COVID-19 a pandemic on 11 March 2020 ☐

