## Supplementary material for "INTERVENTIONS TO IMPROVE PATIENT SAFETY DURING THE COVID-19 PANDEMIC: A SYSTEMATIC REVIEW": Annex E

### Annex E: Risk of Bias of Included Studies

Table 3: Newcastle-Ottawa Scale scoring for cohort studies ^11^

| **Study** | **Selection** | **Comparability** | **Outcome** | **Total Score** | **Assessment** |
| --- | --- | --- | --- | --- | --- |
| **Medication Safety** | | | | | |
| Abdel-Qader, 2022 | 2 | 0 | 3 | 5 | fair |
| Abhisek, 2021 | 2 | 0 | 3 | 5 | fair |
| Besson, 2021 | 1 | 0 | 3 | 4 | fair |
| Perez, 2022 | 3 | 0 | 3 | 6 | fair |
| Prabath, 2023 | 3 | 0 | 2 | 5 | fair |
| Smith, 2021 | 1 | 0 | 3 | 4 | fair |
| Yap, 2023 | 3 | 0 | 1 | 4 | fair |
| **Diagnostic Safety** | | | | | |
| Fattorutto, 2022 | 3 | 0 | 2 | 5 | fair |
| Liu, 2021 | 2 | 0 | 2 | 4 | fair |
| Loveday, 2021 | 2 | 1 | 2 | 5 | fair |
| Scrima, 2020 | 1 | 0 | 2 | 3 | fair |
| Shen, 2022 | 3 | 0 | 1 | 4 | fair |
| Wee, 2020 | 3 | 0 | 2 | 5 | fair |
| Wienhold | 2 | 0 | 2 | 4 | fair |
| **Surgical Safety** | | | | | |
| Ali Hassan, 2020 | 2 | 0 | 2 | 4 | fair |
| Belenje, 2022 | 2 | 0 | 2 | 4 | fair |
| Boffa, 2020 | 2 | 0 | 2 | 4 | fair |
| Bowman, 2023 | 2 | 0 | 2 | 4 | fair |
| Burden, 2021 | 2 | 0 | 3 | 5 | fair |
| Ceraudo, 2021 | 2 | 0 | 3 | 5 | fair |
| Chan, 2023 | 2 | 1 | 3 | 6 | fair |
| Choi, 2020 | 2 | 0 | 3 | 5 | fair |
| Dabek, 2022 | 2 | 0 | 2 | 4 | fair |
| Daniels, 2023 | 3 | 1 | 2 | 6 | fair |
| Erbas, 2021 | 2 | 0 | 2 | 4 | fair |
| Grubbs, 2023 | 2 | 0 | 3 | 5 | fair |
| Habib Bedwani, 2023 | 3 | 0 | 2 | 5 | fair |
| Jiang, 2021 | 2 | 0 | 3 | 5 | fair |
| Joseph, 2021 | 3 | 1 | 3 | 7 | Good |
| Leung, 2021 | 3 | 1 | 3 | 7 | Good |
| Logishetty, 2021 | 2 | 0 | 3 | 5 | fair |
| Monroy-Iglesias, 2021 | 3 | 1 | 3 | 7 | Good |
| Moschovas, 2021 | 2 | 0 | 3 | 5 | fair |
| Narang, 2020 | 3 | 1 | 3 | 7 | Good |
| Pai, 2020 | 2 | 0 | 3 | 5 | fair |
| Ren, 2021 | 2 | 0 | 3 | 5 | fair |
| Sebastian, 2022 | 3 | 1 | 2 | 6 | fair |
| Sivaraj, 2021 | 3 | 0 | 2 | 5 | fair |
| Stagg, 2022 | 2 | 0 | 2 | 4 | fair |
| Turkdogen | 2 | 0 | 3 | 5 | fair |
| Vaidya, 2021 | 1 | 1 | 1 | 3 | fair |
| Wee, 2020 | 3 | 0 | 2 | 5 | fair |
| Wienhold | 2 | 0 | 2 | 4 | fair |
| Yap, 2023 | 3 | 0 | 3 | 6 | fair |
| Zago, 2020 | 2 | 0 | 3 | 5 | fair |
| **Healthcare Associated Infections** | | | | | |
| Gragg, 2021 | 1 | 1 | 2 | 4 | fair |
| Guven, 2018 | 3 | 1 | 2 | 6 | fair |
| Johansen, 2022 | 2 | 1 | 2 | 5 | fair |
| Krauss, 2022 | 2 | 1 | 1 | 4 | fair |
| Li, 2022 | 3 | 1 | 1 | 5 | fair |
| McVey, 2022 | 1 | 1 | 1 | 3 | fair |
| Meda, 2020 | 2 | 1 | 2 | 5 | fair |
| Wee, 2021 | 2 | 1 | 2 | 5 | fair |
| **Pressure Injury** | | | | | |
| Doussot_2020 | 1 | 1 | 1 | 3 | fair |
| Gallagher_2022 | 3 | 1 | 2 | 6 | fair |
| Miguel_2021 | 2 | 1 | 1 | 4 | fair |
| Rana_2021 | 3 | 1 | 2 | 6 | fair |
| **Falls** | | | | | |
| Kwok_2022 | 2 | 1 | 1 | 4 | fair |
| Rana_2021 | 3 | 1 | 2 | 6 | fair |

Table 4: Newcastle-Ottawa Scale scoring for case-control studies ^11^

| **Study** | **Selection** | **Comparability** | **Exposure** | **Total Score** | **Assessment** |
| --- | --- | --- | --- | --- | --- |
| **Medication Safety** | | | | | |
| Ong, 2023 | 2 | 1 | 3 | 6 | fair |

Table 5. Risk of Bias 2 assessment for controlled trials ^10^

| **Study** | **Randomization** | **Deviation from intended intervention** | **Missing outcome data** | **Measurement of the outcome** | **Selection of reported result** | **Overall** |
| --- | --- | --- | --- | --- | --- | --- |
| **Pressure Injury** | | | | | |  |
| Taylor, 2021 | Low | Low | Low | Low | Low | Low |
