## Supplementary material for "INTERVENTIONS TO IMPROVE PATIENT SAFETY DURING THE COVID-19 PANDEMIC: A SYSTEMATIC REVIEW": Annex D

### Annex D: Evidence tables

*Publication language is English unless otherwise noted

**Articles which were hand selected and not part of the literature search results

#### Medication Safety

| **Au, yr** | **Patients, n** | **Country** | **Study design** | **Inclusion criteria** | **Exclusion criteria** | **Description of intervention, innovation, adaptation, or change in practice** | **Narrative Outcome Description** |
| --- | --- | --- | --- | --- | --- | --- | --- |
| Abdel-Qader, 2022 | Not assessed  (110 community pharmacies) | Jordan | Randomized, controlled trial | Pharmacies open 5-6 days/week, at least 2 shifts per day, providing pharmaceutical services for more than 30 patients/ customers per day, connected to stable wi-fi network, adequate pharmacy staff to cover absences of colleagues.    Pharmacy Staff Inclusion:  Ministry of Health license, regular working hours, current WhatsApp user, English and Arabic speaker | Pharmacies newly opened, located more than 7 km from the nearest medical facility (ie, clinics, medical centers), pharmacy manager was the only pharmacy staff    Pharmacy Staff Exclusion:  Pharmacists enrolled in educational program focusing on medication errors (MEs) or prescribing errors (PEs); participation in a study focusing on PEs | Clinical scenarios were developed to provide to pharmacy staff.  Scenarios were transcribed into WhatsApp messages and sent to pharmacists in active group on daily basis over 4 weeks. | - The incidence of Prescribing errors (PE) in COVID-19 patients, PEs per patient, and number of patients with at least one PE reported by participants in the active and control groups were 18.54% versus 3.09% (P = .001), 0.83 versus 0.16 (P = .02), and 2274 versus 595 (P = .01), respectively - The proportions of lethal, serious, and significant errors were 0.74% versus 0.35% (P = .04), 10.52% versus 2.57% (0.002), and 47.88% versus 9.57% (P = .001), respectively - Time taken to detect PEs and the time needed to persuade physicians/patients with PIs between the active and control groups were (3.87 ± 2.78 min) versus (8.29 ± 6.38 min) and (2.03 ± 1.76 min) and (5.02 ± 4.22 min), respectively (all with *P*<.05) |
| Abhisek, 2020 | N/A | Eastern India | Prospective observational, software-based study | Hydroxychloroquine (HCQ) drug-drug interactions (DDI) with other groups of drugs present in the software system | N/A | Comparison Lexicomp software vs Medscape and Drugs.com in assessing Drug Drug Interactions (DDI) of hydroxychloroquine (HCQ) | Total of 279 DDIs of HCQ with individual drugs. Among these, 66.66% were maximum risk rating of C.  Lexicomp software was better than Medscape and Drugs.com in assessing DDIs of HCQ |
| Besson, 2021 | N/A  (two pharmacy compounders - critical care units & four pharmacists) | France  Publication Language: French | Prospective observational | Croix-Rousse hospital (675 beds, only 339 under Rx analysis) | N/A | Intervention included daily updates between hospital pharmacy technicians (PPH) and pharmacists on the status of medication stock and number of prescriptions in progress, including:  1. Disruptions, distributed stocks in different units to have the same presentation in the same department on a given day;  2. Communication by email and/or telephone to physicians and critical care pharmacy executives that are out of supply/available in each unit; creation of a specific table of stock coverage based on consumption over the last seven days was distributed regularly;  ---created multi-professional Hospices Civils de Lyon (HCL) working group bringing together anesthetists-resuscitators, pharmacists, care management to prevent the risks of pharmacy errors related to the delivery of imported drugs not re-labelled by laboratories with re-labelling actions at the central level and drafting of common information sheets.  3. Live communication with doctors and state-certified nurses (IDEs) and doctors by PPH, & “alert” posters, e.g., when introducing new cisatracurium concentrations. | Daily communication between pharmacy, clinicians and nurses on the status of stocks was a key finding.  Calculating the daily per patient quantities to adjust orders and deliveries that was adapted to each unit was an attribute of the intervention.  Due to pharmacists' time restrictions, not all prescriptions were analyzed during this period of crisis leading to a higher risk of errors.  The just-in-time flow of supply and changes in suppliers, when not re-labelled in French, justifies the presence of compounders and pharmacists in the care units, 7 days a week.  The use of ‘secure’ cabinets do not provide safety medication management compared to assigning to compounders in each unit (as opposed to 24/7 coverage). |
| Perez, 2022 | 438 | France | Prospective cohort study | All patients admitted consecutively over 1 month to COVID-19 units and who had received from pharmaceutical analysis | Patients who had not received pharmaceutical analysis, unknown COVID-19 status | Medical prescriptions were analyzed daily throughout hospital stay by three clinical pharmacists.  When a trained pharmacist detected an opportunity to improve care, a pharmaceutical intervention (PI) was launched. | A total of 188 pharmaceutical interventions (PIs) were performed on the medication prescriptions of 118 patients: 64 and 54 patients for positive and negative groups, respectively (p=0.236), resulting in an average PI rate of 1.6 PIs/patient  Physicians’ acceptance rate of PIs for COVID-19- 19- positive patients was 88.5% (92/104), and that for COVID-19- 19- negative patients was 90.5% (76/84) |
| Prabath, 2022 | 135 | India | quasi-experimental pretest-posttest | Undergraduate nursing students | students unavailable at the time of circulation of the questionnaire were excluded | Participants completed a validated knowledge, attitude, and practice (KAP) questionnaire through an online messaging software pre-intervention. Following the questionnaire, an educational presentation was conducted for 45-60 on aspects of adverse drug reaction (ADR) reporting.  Aspects included: the definition of ADR, classification of ADR, the necessity of ADR reporting, functions of Pharmacovigilance Programme of India (PvPI) and ADR Monitoring Centre, ADR reporting methods, how to fill an ADR reporting form, and role of healthcare professionals in pharmacovigilance taught to the students.  This was followed by a post-test questionnaire on the same day. | - More than 75% of participants acquired knowledge about adverse drug reaction (ADR) Monitoring Centre and Pharmacovigilance Programme of India (PvPI). - More than 30% increase in awareness about eligible individuals and different methods of ADR reporting, a suspected medication that can be reported and potential benefits of pharmacovigilance (PV) practices were observed. - More than 50% of participants had observed ADR during their clinical postings but less than 50% reported ADR. |
| Smith, 2021 | 527,471 | USA | Simulation | Commercial or Medicare Regence health plan  On two or more medications | Participants with no drug claims data | A simulation strategy based on medication risk score could be used as a blueprint to estimate adverse drug events (ADE) risk with repurposed drugs. All repurposed drugs studied were associated with an increased risk of ADEs Mean 47 (0-89) | - The addition of all repurposed drugs increased drug-induced Long QT Syndrome and increased the medication risk score (MRS) (median increased by 2 to 7 points; p < 0.001 - The number of patients at moderate risk enhanced by ~ 60% to 420% compared to baseline for the commercial insured group (n = 10,028 2.3%) and by~ 30% to 200% for the Medicare group (n=6726,7.3%) - The number of patients at high risk increased by ~ 90% to 570% compared to baseline for the commercial insured group (n=5633, 1.3%) and by 70% to 310% Medicare groups (n=4815, 5.2%) |
| Yap, 2023 | 77 | Singapore | prospective comparator | - age ≥5 years - abdominal pain duration ≤48 h - ultrasound (US)/Computer Tomography scan confirmation of Acute Uncomplicated Appendicitis (AUA) - US appendiceal diameter 6–11 mm with no features of perforation/abscess collection and no faecolith | - patients with generalized peritonitis on clinical examination - patients with a significant medical history or concurrent medical condition that may potentially place them at a higher risk for complications | Eligible patients were counselled regarding both treatment options – upfront surgery versus non-operative treatment (NOT) with antibiotics alone.   - For patients managed with NOT, initial intravenous (IV) antibiotics, were administered for a minimum of 24 h. Patients were discharged home with a course of oral antibiotics for 10 days. On discharge, patients were given a diary to record their home recovery. - For patients managed through laparoscopic appendectomy, received IV antibiotics for 24 hours perioperatively. Patients were discharged when able to resume normal diet and were without early post-operative complications. | Success of non-operative treatment (NOT) at index admission was 90.7% (39/43).  NOT failure rate at 27 months’ follow-up was 37.2% (16/43).  Of the NOT failures, 1 appendix was normal on histology while only 1 was perforated.  There were no significant differences in secondary outcomes between both groups except for length of stay (LOS) of late NOT failure. |

#### HAI

Table 1. Population Characteristics

| **Au, yr** | **Patients, n** | **Country** | **Study design** | **Inclusion criteria** | **Exclusion criteria** | **Description of intervention, innovation, adaptation, or change in practice** | **Narrative Outcome Description** |
| --- | --- | --- | --- | --- | --- | --- | --- |
| Gellert, 2022 | N/A | USA/UK | Implementation | Hospital/Health systems implementing an identity access and management (IAM) and single sign on (SSO) technology | N/A | Leveraging identity access and management (IAM) and single sign on (SSO) technologies to improve clinical, infection control and/or operational workflows.  Eight innovative/valuable hospital use cases are described: symptom-free attestation by clinicians at shift start; detection of clinician exposure/contact tracing; reporting of clinician temperature checks; inpatient telehealth consults in isolation units; virtual visits between isolated patients and families; touchless single sign-on authentication; secure access enabled for rapid expansion of personnel working remotely; and monitoring of temporary worker attendance. | Creating partnerships between hospitals and their information technology (IT) vendors can help implementation of innovative solutions.  Described use cases provide justification for an ‘innovation sandbox’ through which care delivery organisations can explore and innovate needed functionality, adding value and impact to existing products/services. In today’s cost-conscious performance-focused healthcare environment. |
| Gragg, 2021 | N/A | USA | Observational | Emergency department | N/A | An eight-pronged response plan was developed and implemented within an emergency department to enhance patient and staff safety during the pandemic.   1. Parallel Emergency Department (ED) Lanes: redesigned ED floorplan to create a Respiratory ED and a Medical/Trauma ED for patients with non-respiratory conditions. 2. Universal respiratory precautions: use of personal protective equipment (PPE) for airborne, droplet and contact precautions 3. Respiratory Drive Through (RDT) for high volume evaluation of patients within their vehicles 4. Area Support Medical Company (ASMC) equipped with a tent-based treatment facility 5. Provider Triage to rapidly evaluate patients who bypass the RDT/ASMC and access the ED directly 6. ED Quarterback Patient Liaison to assist patients with accessing their primary care provider (PCP) for concerns that are better handled at their primary care center. 7. Virtual Registration to minimize contact between patients and ED administrators and decrease overall ED length of stay 8. Virtual Ward to monitor patients remotely with cardiac and pulmonary telemetry to discharge COVID-19 ED patients that require monitoring but not immediate intervention | The implementation of the COVID-19 response plan resulted in the ability to test over 20,000 patients for COVID-19 with zero cases of nosocomial COVID-19 infections among health care workers. |
| Guven, 2021 | 402 | Turkey | Retrospective observational | Patients hospitalized for ≥ 48 hrs. | Patients hospitalized for imaging and one-day chemotherapy discharged on the same day of hospitalization | Isolation and hygiene measures were implemented in an oncology hospital at the start of the COVID-19 pandemic which include:   - placement of warning signs for social distancing, - the use of masks by the health care workers, - symptom screening and triage - increased use of telemedicine, - inpatient visitor restrictions, - efforts for a shorter hospital stay, and - hospitalization of patients with respiratory symptoms in isolated rooms. | - Intervention demonstrated a significant reduction in overall nosocomial infections in the first 3 months of the pandemic compared to the previous year (18.6% vs 32.2%), corresponding to a decrease in different types of healthcare associated infections (HAIs), namely pneumonia, urinary tract infection (UTI), bacteremia and intraabdominal infections. - Preventive measures for COVID-19, led by improved hygiene and distancing measures for health care workers and visitors, increased use of telemedicine and adaptations for early discharge could lead to a reduction in nosocomial infection rates in oncology wards. |
| Johansen, 2022 | 43,755 | USA | Retrospective cohort | Medicare Part B claims submitted by outpatient dialysis facilities:  -Patients undergoing full-care, in-facility hemodialysis with a central venous catheter during the last 7 days of each calendar month.  -Intravenous administration of antibiotics without modifier code AY, which indicates treatment unrelated to kidney failure.  -Any hospitalization with a discharge diagnosis of infection related to a catheter. | N/A | Infection-control measures were implemented to reduce transmission of COVID-19 within dialysis centers at the start of the pandemic which include:   - Use of personal protective equipment (PPE) - Frequent hand hygiene - Frequent cleaning and disinfection of surfaces and dialysis machines and stations | - Results showed a decreased rate of antibiotic administration and catheter associated bloodstream infections during the first 6 months of the COVID-19 pandemic compared to pre-pandemic rates (between 20%-21% and 17%-24% lower, respectively). - These findings suggest that infection control measures implemented in dialysis facilities to reduce transmission of COVID-19 such as gowning, masking, and increased disinfection may reduce catheter-associated bloodstream infections in hemodialysis patients. |
| **Krauss, 2022 | N/A (49 ICU teams) | USA | Quality improvement | Intensive care units (ICUs) with elevated rates of central line-associated bloodstream infections (CLABSI) and/or catheter-associated urinary tract infections (CAUTI) participating in the Agency for Healthcare Research and Quality Safety Programme for ICUs: Preventing CLABSI and CAUTI | N/A | With a focus on a series of 30 min, semistructured, retrospective exit interviews held between members of the national programme team (NPT) and 4 staff members, which represented 11 of the participating units with the aim of obtaining a holistic perspective from the units on the programme and their experience in it.  Reviewed responses from semistructured exit interviews, self-assessments on healthcare- associated infection (HAI) prevention activities, participant-created action plans, chat-box discussions during webinars and informal correspondence. | Between 80% and 90% of units adopted principles such as obtaining the support of senior leadership and identifying nurse and/or physician champions.   1. Considering the pandemic, other intensive care units (ICUs) were able to find ways to flex key activities, such as their daily huddles. 2. Participants shared that multidisciplinary rounds were modified but still maintained to accomplish established protocols, such as discussion of line necessity. 3. Participants reported that flexibility was key for teams to find ways to apply principles and adapt best practices amid pandemic challenges. This became particularly clear through the revised action plans and other materials submitted during the pandemic. Unit goals were the most frequently changed section in revised action plans, with 20 of 49 participating units reporting changes. 4. Continued buy-in from teams engaged in Infection Prevention (IP) activities is an important component. Respondents to exit interviews reported higher team cohesiveness and commitment than they experienced prior to the pandemic. |
| Li, 2020 | 269 | China | Retrospective observational | Patients who have received catheter ablation for symptomatic arrhythmia with poor response | Patients who received a catheter ablation but have adequate response | Creation of a modified workflow for arrhythmia disease catheter ablation was established, which was agreed upon among the local cardiologists and was comprehensively evaluated within the risk–return ratio for both patients and health care workers.  - All patients admitted to hospital were required to undergo risk assessment for COVID-19‐19‐related symptoms.  -Once symptoms or contact was confirmed, patients were marked by risk. High risk patients were transferred to a COVID-19 designated hospital for quarantine and testing.  Low-risk were divided into urgent and emergent cases, semi-urgent cases or non-urgent/elective cases.  - For urgent or emergent cases, there were a ‘green channel’ and once excluded from high‐risk, they immediately received routine blood tests, lung computed tomography (CT), and SARS‐CoV‐2 nucleic acid detection. Suspected and confirmed cases were immediately transferred to a COVID-19‐19 designated hospital for treatment. For other urgent cases, catheter ablation was performed in a special catheterization laboratory dedicated to emergency interventional surgeries. | Prevention measures ensured that catheter ablation may be safely performed with an infection rate of 0% at the study’s centre;  a beneficial clinical outcome of catheter ablation can be achieved regardless of the type of arrhythmia, with 95.9% patients becoming free of arrhythmias (±AAD);  theEuropean Heart Rhythm Association (EHRA) score significantly improved following ablation in atrial fibrillation (AF) patients; and  the rate of major complications was 0%. |
| McVey, 2022 | Not applicable | USA | Prospective longitudinal | A tracked event (e.g., central line-associated bloodstream infections (CLABSI) and/or catheter-associated urinary tract infections (CAUTI)) had occurred on a unit within the past year | N/A | The Nurse- Sensitive Indicator Quality Improvement (NSIQI) Toolkit for CLABSI and CAUTI prevention was adapted from K-cards and other best practices for HAI reductions and comprised of 2 primary tools:   - Zero Boards, to track and visualize data trends on patient safety indicators. Updated daily by assigned nurse. - Report Cards, to track nurse driven CLABSI and CAUTI bundle elements. Completed each shift, assigning a letter grade based on central line and Foley catheter maintenance audits.   Leadership rounds were included to support toolkit implementation on each unit: daily for two weeks, followed by weekly for one month, followed by monthly rounds on each unit. | CLABSI and CAUTI standardized infection ratio (SIR) data were collected on nursing units at baseline and post intervention, and showed a reduction in hospital-wide CLABSI and CAUTI rates during the 10-month post-intervention period (19% and 19.4% reduction in CLABSI and CAUTI SIR, respectively).  These findings suggest that implementation of the NSIQI toolkit may help reduce and sustain CLABSI and CAUTI infection rates during surges in the pandemic. |
| Meda, 2020 | A 12-bedded critical care | UK | Prospective cohort/quality improvement | Patients and healthcare workers (HCWs) with a 12-bed critical care unit. | N/A | - Created a monitored hand hygiene protocol. - Sampled near-patient environment: found increased gram-negative bacteria (GNB) on surfaces. - Discussed with critical care health care workers (MDT and hospital senior mgt), and changed PPE from long-sleeved to short-sleeved gowns. | - Use of a short-sleeved gown increased hand hygiene compliance, enhanced cleaning/disinfection of the surfaces, and CLABSI reduced from 3 to 0 post intervention. - 11.5% of surfaces contaminated with GNB down to zero post-enhanced cleaning intervention. |
| Pai, 2020 | 593 | USA | Prospective cohort/quality improvement | All pre-operative patients | N/A | A multidisciplinary quality improvement team was formed.  The existing preoperative evaluation (POE) clinic was utilized as a centralized method to provide COVID-19 testing, symptom screening, and infection prevention education in addition to routine preoperative medical optimization.  -During POE triaging calling, nurses provided education on isolation, personal hygiene, and face mask use.  -Patients with symptoms but testing negative for COVID-19 were referred to a COVID-19 Virtual Clinic (CVC) for further evaluation  -If patients tested positive for COVID-19, they were referred to the CVC for telephone or video follow-up and their surgeries were postponed.  DMAIC (define, measure, analyze, improve, control) methodology was used and the study was limited to analyzing scheduling and administrative data. | With the new process, the percentage of patients with COVID-19 testing results returned before surgery increased from 10% to 100%.  Emergent surgeries in which physicians felt a delay in waiting for COVID-19 test results would adversely affect their well-being, would proceed as if they had tested positive.  Of the 593, only 2 patients were found positive during preoperative testing and had their surgeries rescheduled. |
| Pate 2022 | 874-bed, level 1 trauma and academic  medical center | USA | Prospective cohort/quality improvement | Select surgical and oncology units of the hospital in Charlotte, NC. | Critical care and maternity units | Implement audits with a CLABSI rounding team supported by master’s prepared nursing roles, infection preventionists | When they reduced the audits during the COVID-19 surges, there was an increase in CLABSIs. There is no description or any focus on how to adapt the basic infection prevention practices during the COVID-19 surges. |
| **Wee, 2021 | N/A (1,785 bed hospital) | Singapore | Implementation | All HAIs during study period, February 1, 2020-August 31, 2020 | HAI rates outside study period | A campus-wide multimodal Infection Prevention and Control (IPC) bundle was implemented.  Patients with respiratory symptoms but no epidemiological risk factors for COVID-19 were segregated in designated clinical areas (termed as respiratory surveillance wards [RSWs]).  Beds were distanced to at least 1.5m to encourage safe distancing. Usage of surgical masks among hospitalized patients was mandatory.  Confirmed COVID-19 cases were housed in dedicated airborne-infection-isolation-rooms (AIIRs), either in the SGH's purpose-built 51-bedded isolation ward (IW) or in a 50-bedded IW extension containing AIIRs modified from containers that was constructed during the COVID-19 pandemic  There were also enhanced campus-wide IPC measures were also introduced in the general ward setting, to mitigate the potential risk of an unsuspected case presenting outside of areas designated for COVID-19 management.   - universal masking policy for all HCWs in clinical areas was introduced, with usage of a surgical mask the mandatory minimum - Regular hand hygiene with alcohol handrub was also re-emphasized - prepandemic all patient areas were cleaned with 1:1000 hypochlorite-based disinfectant, at a frequency of at least 3 times a day; during the pandemic, cleaning practices were reinforced and regular environmental cleaning audit using fluorescent markers (Glogerm) was maintained - cohorting was maintained in the general ward setting for patients with muti-drug resistant organisims (MDROs) - additional isolation capacity was provided through the container extension - a no-visitor policy was enforced throughout hospital in tandem with the community-wide imposition of an elevated set of safe-distancing measures | Rates of HAI over a 7-month period during the COVID-19 pandemic (February 1, 2020-August 31, 2020) after the introduction of enhanced IPC measures were compared with rates of HAI over the preceding 2 years (January 2018-January 2020).   - Over the corresponding period, the hospital saw ≥1,600 cases of COVID-19, with no evidence of patient-HCW transmission.   After introduction of enhanced IPC measures:   - The incidence of PCR-proven health care-associated respiratory-viral-infection (HA-RVI) was 0.83 cases per 10,000 patient-days (22 cases; 264,904 patient-days) - The incidence rate ratio (IRR) of PCR-proven HA-RVI per 10,000 patient-days between the 2 periods (pre- and post-pandemic) was 0.08   During the pandemic:   - admissions for community-acquired respiratory-viral-infections (RVIs) at our institution remained stable. - For enveloped HA-RVI, the rates fell from 6.05 cases per 10,000 patient-days (618 cases) to 0.45 cases per 10,000 patient-days (12 cases) after the introduction of enhanced IPC measures - For nonenveloped HA-RVI, the rates fell from 3.63 cases per 10,000 patient-days to 0.38 cases per 10,000 patient-days (IRR = 0.10) |

#### Diagnostic Safety

| **Au, yr** | **Patients, n** | **Country** | **Study design** | **Inclusion criteria** | **Exclusion criteria** | **Description of intervention, innovation, adaptation, or change in practice** | **Narrative Outcome Description** |
| --- | --- | --- | --- | --- | --- | --- | --- |
| Fattorutto, 2022 | 32 | Belgium | Retrospective implementation | Adults (18 years and older)  Patients admitted to the ICU  Patients with acute hypoxemic respiratory failure characterized by a PaO_2_/FiO_2_ ratio ≤ 150 mm Hg  Patients with confirmed COVID-19 infection | Only the first admission was retained for patients admitted more than once  Patients with more than one stay who were readmitted to the ICU. | This study created a thromboprophylaxis algorithm for patients with severe COVID-19.  The control group included patients who were treated before April 9, 2020, and received standard or boosted prophylaxis with low molecular weight heparin (LMWH).   - The standard prophylactic dose of LMWH corresponded to enoxaparin 4000 IU daily, subcutaneous (s.c.) if body weight (bw) was < 100 kg or 6000 IU daily, s.c. if bw was > 100 kg. - If the patient was placed on invasive mechanical ventilation or high flow oxygen therapy, boosted prophylaxis was prescribed (according to bw) twice daily (b.i.d.). - After April 9, 2020, all patients had targeted management of heparin therapy based on the algorithm. - Patients treated exclusively according to our algorithm were the protocol group | - Safety: the implementation of the algorithm did not increase major bleeding rate - Efficacy: Results were consistent with observations from other studies focused on thromboembolism (TE) in COVID-19 ICU patients. The use of high‐intensity prophylaxis treatment (enoxaparin 40 mg b.i.d. in 75 patients) was associated with a lower incidence of TE (12.2%), without increasing major bleeding. - Biological measurements: antithrombin III (ATIII) activity was decreased well below the lower limit of normality which was close to the findings from various studies. The MCF- EXTEM™ assays were outside the upper limit of normality and the amplification of fibrinogen and D-dimer levels were well beyond the limits of normality. |
| **Liu, 2021 | 4,589 (132 clinicians) | China | Retrospective | All patients who received remote diagnosis and treatment via online consultation services | Not defined | To describe features and clinical symptoms of patients who receive remote diagnosis and treatment.  An online survey was administered to the outpatient consultation services and included clinicians from the Department of Infectious Diseases, the Respiratory Department, the Department of Critical Care Medicine, and the Department of Psychology and Psychiatry.  Clinicians were divided into 5 teams:   - 2 teams provided consultation services for adult patients (adult team) - 1 team for pediatric patients (pediatric team) - 2 teams for patients with psychological problems (psychological team)   Patients can scan the official QR (Quick Response) code or follow the WeChat public account to consult with a physician. The account will direct them to the expert consultation interface and allows access to the online outpatient consultation. Patients can interact with the clinicians online via voice, text, photo, and video.  At the end of the remote consultation, the platform will automatically open an electronic questionnaire, which the patients have the option to fill. The questionnaire includes star ratings and open answers. A minimum of 1 star represents high dissatisfaction, and a maximum of 5 stars represents high satisfaction. Four stars and above represent satisfaction. Following this, patients can input their own opinions and suggestions. | By the end of the study period, the 5 expert teams had provided remote consultation services for 4589 patients   - the 2 adult teams provided consultations to 4399 adult patients, accounting for 95.86% of the sample - the pediatric team provided consultations to 80 (1.74%) pediatric patients - the 2 psychological teams provided psychological counseling for 110 (2.40%) patients   The most common symptom that patients initiated a consultation for was fever (n=2383 patients), followed by cough (n=1740), nasal obstruction (n=794 patients), fatigue (n=503), and diarrhea (n=276).  Among the 110 patients who received psychological counseling, 7 patients reported psychological stress and anxiety due to their history of epidemiological exposure.  985 patients responded to the satisfaction questionnaire, of whom 98.1% (n=966) were satisfied with the service (rated as 4 stars or above).  In the 110 cases of psychological counseling, 47 cases provided feedback, with satisfaction ratings greater than 4 stars was 100%. |
| Loveday, 2020 | N/A (Various depending on group modeled) | UK | Modeling study | Patients in the colorectal cancer (CRC) pathway to diagnosis and treatment in age-specific and stage-specific strata (stage 1, 2, 3)  Subset of urgent symptomatic patients (2-week wait [2WW] for CRC diagnostic referral) | Stage 4 cancers | Baseline survival and life years lost were modeled as a result of delays of 2-6 months in the urgent diagnostic pathway, given no fecal immunochemical testing (FIT) triage.  The study modeled FIT triage in the urgent symptomatic population to mitigate survival decrement from 2-, 4- and 6-month delays in diagnostic pathway during COVID-19 disruptions related to shutdowns of endoscopic services. | - Adopting a FIT threshold of 10 µg Hb/g faeces would reduce immediate colonoscopy requirements in the symptomatic group to 18% of normal (92 051/511 394), and identify 89% (9991/11 226) of CRCs. - While adopting a higher FIT thresholds may reduce immediate colonoscopy requirements, it is at the expense of an additional 150/326/517 deaths per year if background delay rate is 2/4/6 months.   A cut- off of 2 µg Hb/g offers improved sensitivity (96%, 10 777/11 226) but only reduces colonoscopies to 37% of normal rates (189 216/511 394).” |
| Scrima, 2020 | 46 | Italy | Prospective observational | Patients meeting clinical stratification criteria for nuclear testing | Patients with suspicious symptoms for COVID-19 | Rapid design of a new safety protocol for goals of continuous patient care during pandemic, protecting both health care professionals and patients without lengthening the waiting lists, and better select patients to observe incidence of inducible ischemia   - The new ad hoc protocol allowed the unit to continue safe performance of stress myocardial perfusion scintigraphy. - Designed to ensure least contact between patient and health care professionals for the shortest time. - Replaced the standard exercise-induced stress test with the pharmacological test using regadenoson. | - Strict ad hoc hygiene protocol avoids diagnostic-therapeutic delay and lengthening of waiting lists. - None evidence of complications. - Short follow-up of patients discharged was carried by phone and no complaints were registered as well as absence of COVID-19 infection symptoms. - No healthcare worker infections occurred during the study period |
| Shen, 2022 | N/A – 14,230 safety reports | USA | Computerized and manual review of patient safety event reports | All event reports | N/A | Safety event reports during the study period were evaluated using two complemetnary pathways.  1. Reports with explicit mention of COVID-19;  2. All reports without explicit mention of COVID-19 where natural language processing [NLP] plus logic-based stratification was applied to identify potential cases  Cases were evaluated manually to identify diagnostic error/delay and categorize error type using a recently proposed classification framework of eight categories of pandemic-related diagnostic errors.  1. Classic  2. Atypical COVID-19  3. Anchor  4. Secondary  5. Delayed Presentation- Acute Condition  6. Delayed Presentation – Chronic Condition  7. Strain  8. Unintended | Each workflows identified a set of diagnostic error incident reports for further review and categorization.  Of 14,230 reports, 95 (0.7%) were identified as cases of diagnostic error/delay.  Pathway 1 (n = 1,780 eligible reports) yielded 45 reports with diagnostic error/delay (positive predictive value [PPV] = 2.5%), of which 35.6% (16/45) were attributed to pandemic-related strain. In  Pathway 2, the NLP–based algorithm flagged 110 safety reports for manual review from 12,450 eligible reports. Of these, 50 reports had diagnostic error/delay (PPV = 45.5%); 94.0% (47/50) were related to strain. |
| Wee, 2020 | 8,437 | Singapore | Prospective observational | Patients admitted to surgical department | N/A | Risk-stratification strategy to ensure appropriate infection prevention measures for all potential COVID-19–infected patients requiring operations, to ensure safe delivery of surgical services | The risk-stratification procedure allowed continuity in surgical services without testing all admissions.  Details of the diagnostic and therapeutic care were described for the six (6) COVID-19 detected cases identified by the risk-stratification and infectious control triage protocols. |
| Wienhold, 2021 | 111 | Germany | Implementation | - ≥18 years of age - have been assigned an ASA I/II classification based on the ASA classification system - have elective surgery scheduled at least 14 days in the future - have access to a Windows 10-based personal computer with a webcam | - language or cognitive barriers - high-risk procedures with need for postoperative ICU stay - acute infection with required auscultation | Patients were assigned to receive teleconsultation for preoperative evaluation and to complete a subsequent survey.  Patients received an individual username, password and download link for software TARA (Telemedical Anamnesis and Risk Assessment)  Patients were also asked to complete study questionnaires, based on a previous study ^86^ . | Most anaesthetists rated telepreoperative evaluation as equivalent to an on-site meeting (93.7%), and expressed interest in continuing the practice in the future (98.0%).  94.8% of anaesthetists claimed that they were able to assess the individual perioperative risk during telepreoperative evaluation.  While 57% of patients already had an on-site preoperative evaluation for other reasons, most patients considered telepreoperative evaluation as convenient as an on-site meeting (98.2%). |

#### Surgical Safety

| **Au, yr** | **Patients, n** | **Country** | **Study design** | **Inclusion criteria** | **Exclusion criteria** | **Description of intervention, innovation, adaptation, or change in practice** | **Narrative Outcome Description** |
| --- | --- | --- | --- | --- | --- | --- | --- |
| Ali Hassan, 2020 | 28 | Canada | Cluster | Candidates for coronary artery bypass grafting | Not stated | Utilization of a nonsealed endoscopic vessel harvesting (EVH) approach during coronary artery bypass surgery. | Of the 28 patients, 23 patients under- went endoscopic harvesting of the great saphenous vein and 5 underwent endoscopic radial artery harvesting.  This approach does not necessitate the use of CO2 insufflation and thus mitigates the risk of possible viral aerosolization. |
| Belenje, 2022 | 200 | India | Prospective | Elective surgeries | Patient not undergoing surgery | Implementation and outcomes of guidelines were formulated in-house and outreach was done across other facilities to support urban and rural Telangana state in providing effective screening services while implementing the new guidelines.  Delay in follow-up of low-risk staged retinopathy of prematurity (ROP) by 1 or 2 weeks and laser treated zone 1 ROP by 1 week more than the old guidelines in low-risk babies decreased the frequency of follow up by at least 2 visits per baby.  Treatment of the disease on the same day showed a decrease in the frequency of follow up visits and likelihood of presentation with threshold ROP due to missing the follow-up visits. | 106 (53%) infants who were low risk while 94 (47%) infants with high risk were followed up as per the old guidelines.  Out of the 106 infants (212 eyes) managed by the new guidelines, good outcome (group 1) was seen in 102 (96.2%) infants. Twenty seven of the 102 infants had some form of ROP and 5 of these infants needed treatment. |
| Boffa, 2020 | 122 | USA | Prospective | Confirmed or suspected cancer, whose clinical scenario met the current departmental criteria for appropriateness of surgery (i.e., ‘‘urgent’’ status) were eligible | Those patients not meeting clinical scenarios | COVID-19-minimal surgical pathway was conducted to the risk of hospital-associated COVID-19 infection by preventing patients on the pathway from encountering people, places or materials: Preoperative visits to the hospital were minimized and initial consultations were via telemedicine. Best practices were implemented on preventing COVID-19.  Once past the initial screening station at the entrance of the hospital, patients were directed through the hospital using a pathway to mitigate exposure.  -They would be guided through admitting, preoperative holding, operating rooms, recovery area, and inpatient units ensuring they were physically removed from any possible areas used for confirmed or suspected COVID-19 infections (clustering pathway patients). | - No postoperative COVID-19 infections - Nine cases were seen in emergency department and eight cases were admitted - While the surgical pathway was neither novel nor innovative, it was implementation of best practices and a common-sense approach to isolating noninfected patients from infected patients. |
| Bowman, 2023 | 15 | USA | Retrospective cohort | Patients who underwent same-day elective colon resections were selected based on:   - preoperative health assessment - current medications - risk for loss to follow-up - assistance at home - distance from hospital - type of surgery | Patients with American Society of Anesthesiologists (ASA) physical status class 3 or higher | Having Outpatient Major Elective (HOME) – Robotic Colon Resection Protocol was followed.  Patients who met operating room and post-anesthesia care unit (PACU) criteria were discharged same day from the recovery unit.   - Intraoperative criteria:   Remaining hemodynamically stable without need for vasopressor support or blood transfusion  Estimated blood loss <150mL  Case duration < 4 hours  -Excludes cases converted to open   - PACU criteria:   - Afebrile, normotensive and non-tachycardic   - Able to void   - Pain controlled   - No nausea/emesis   - Observation for minimum of 3 hours | - A total of 73% of patients undergoing same-day discharge. - There were no readmissions or complications during the perioperative 30-day period. |
| Burden, 2021 | 128 | UK | Prospective | Level 1a, 1b, 2, and 3 surgical cases – successive 31-day enrollment | Patients where priority level principles did not apply or were qualified for surgery | Assessment of clinical need was determined using priority level principles and applied to cases to define the urgency:   - Priority level 1a (emergency surgery required within 24 hours) and 1b (urgent surgery required within 72 hours) were planned to be delivered at the RDEFT. - All other cases for which a delay to treatment would result in harm or excess mortality were identified. - Complex surgical cases likely to require intensive or high-dependency postoperative care were allocated to be performed at RDEFT where higher-care facilities are available.   Strategies for delivering time-critical surgical services and prioritizing patients for surgery:  For all priority level 2 and 3 patients, specialties were recommended to consider the following factors when prioritising patients for surgery:  • scheduling priority according to likelihood of surgery being curative: this assigns one of six levels for the proposed treatment, with levels 1–4 being curative intent and priority level 1 having greater than 50% chance of success8  • alternative treatments: the presence or absence of equivalent and available non-surgical treatment options  • frailty score: characterised using the Rockwood Frailty Scale20  • COVID-19 vulnerability level: this is a scale from 1 to 3 comparing expected excess mortality in the event of contracting COVID-19 to that of a fit 70-year-old person18  • anticipated length of hospital stay, intensive care admission and other special requirements.  A model of care delivery for a clean-site along with its sustainment:   - COVID-19 testing requirement for patients undergoing elective surgery - National guidance on donning and doffing from Public Health England was followed - Inline filters were used for insufflation and desufflation | During a 31-day period, 128 surgical procedures were performed. Of those tested in the 14-day period following surgery, none tested positive for COVID-19.  Complications were rare and none was severe  Readmission rate was 1.56%. |
| Ceraudo, 2021 | 119 | Italy | Retrospective implementation | All admitted patients to the neurosurgical ward | Not defined | This study developed preventive measures to limit any possibility of COVID-19 spread, according to the principles of epidemiologic prevention and suggestions from recent literature.  A crisis unit commission for the management of COVID-19 was created. Guidelines were developed to maintain high-quality health care and security for both patients and HCW. Additionally, 2 level of PPE were established, primary and secondary. | No cases of positivity for severe acute respiratory syndrome coronavirus-2 infection were found, and no surgical cases were postponed. |
| Chan, 2023 | 645 | UK | Retrospective comparison | Patients admitted from primary care  Self-presentation to Emergency  Tertiary transfers and inpatient referral transfer to surgical care | Not defined | In addition to COVID-19 testing status and imaging data, factors that contribute to disease severity like:   - duration of symptoms prior to presentation, admission clinical early warning scores (EWS) - initial blood tests - microbiology cultures - radiology tests   The Charlson Comorbidity Index (CCI) was calculated as a measure of pre-existing comorbidity.  Outcomes including readmissions and deaths within 30 days of discharge were also recorded. | When comparing the results to the same period in 2019, they found:   - a 50.0% reduction in admissions - no statistical difference between groups for patient age, sex, body mass index (BMI) or CCI - proportion of mixed/multiple ethnic groups, Asian, Black and Other patients admitted during the COVID-19 period was 20.6% lower compared to non-COVID-19 period. - No statistical difference in admissions by socio-economic status |
| Choi, 2020 | 13 | Korea | Retrospective | Patients with MERS and COVID-19 who underwent tracheostomy | Not defined | Tracheostomy protocol consisted of:   - enhanced PPE (coverall clothing including head cover, shoe covers, two pairs of surgical gloves, powered air purifying respirators (PAPRs) and N95 respirators)   -primary surgeons and assistants also used an outer surgical gown and gloves   - simplified procedures (no limitation in the use of electrocautery and wound suction, no stay suture, and delayed cannula change) - use of a validated screening strategy for healthcare workers - creation of a designated COVID-19 tracheostomy team comprised of one highly experienced head and neck surgeon, two attending ICU specialist (one to manage ventilator/endotracheal tube, one to assist with the procedures) and a senior ICU nurse | No cases of transmission of MERS nor COVID-19 occurred |
| Dabek, 2022 | 76 | Ukraine/USA | Prospective | Candidates for surgical intervention with considerations for:   - Functional deficit - Acuity of need - Indicated surgical procedure was required - Need for multiple procedures - Co-morbidities | Not defined | Local providers assisted with pre-selection of candidates by reaching out to patients’ families and colleagues.  Families were interviewed and examined via Skype, Zoom or Viber. Attempts were made to select patients requiring timely intervention, or those who would receive the highest benefit from a procedure.  In addition to risk assessments and selection of patients, modified follow ups (via telemedicine) and outcomes were recorded.  Traditional outreach clinics were cancelled during this study to reduce the risk of COVID-19 exposure. This was replaced with the use of telemedicine screening.  Education and training was an essential component of the missions. During selection of foreign staff and volunteers, qualifications and skills were screened by team members and trainings sessions occurred through Zoom. Ultimately 12 clinicians and residents and six nurses were selected. Staff were trained on case setup, instruments, equipment and proper sterile techniques. Didactic teaching was also incorporated into trainings.  Implementation of the World Health Organization preoperative checklist was incorporated to reduce the risk of perioperative complications. | Most common reconstructive needs included local tissue rearrangement (92%), scar release and skin grafting (72%), extremity reconstruction (60%), facial reconstruction (28%), and breast reconstruction (4%).  No postoperative complications were reported at 2-months follow up.  No patients reported COVID-19 symptoms during outreach or within 1-month follow up.  No staff experienced COVID-19 symptoms. |
| Daniels, 2023 | 274 | UK | Prospective cohort | - Pre-operative patients over16 years - Patients attending THMC for ENT outpatient clinic - Patients with nasal/nasopharyngeal swab for COVID-19 as part of their normal care - Able to provide informed consent | - Adults unable to understand study information and provide consent - Adults who needed immediate hospitalization | Questionnaires were administered to patients and staff to assess the acceptance of point of care testing (POCT). | Most respondents agreed or strongly agreed that POCT:   - improved care management (79.7%) - decreased administrative time (65.8%) - reduced the risk of canceled appointments (74.7%) - traveling time to do COVID-19 test (91.1%) |
| Erbas, 2021 | 24 | Turkey | Retrospective | - Patients aged over 18 years - Diagnosed with COVID-19 - Requiring percutaneous tracheostomy | - Patients under 18 years - Refusal to participate | Use of an aerosol box during tracheostomy procedure on patients with COVID-19 | - Four patients experienced minor bleeding related to performing the percutaneous tracheostomy using the aerosol box. - No healthcare workers were infected with SARS-CoV-2 while performing the tracheostomy procedure. - Use of the aerosol box has adequate efficacy and safety when performing a percutaneous tracheostomy |
| Grubbs, 2023 | 89 | USA | Prospective | Low risk for outpatient surgery | Patients who were moderate or high-risk functional status and without psychosocial support | Creation of a pathway focused on detecting and preventing early post-operative complications when conducting outpatient bariatric surgery.  The evaluation tool used the Obesity Surgery Mortality Risk score in addition to assessment of comorbidities and functional status. The Obesity Surgery Mortality Risk Score is a validated assessment that predicts mortality of patients undergoing bariatric surgery  The tool primarily used assessment of comorbidities and Obesity Surgery Mortality Risk score to identify low-risk patients.  Patients with a score of 0–1 were generally placed in a low-risk group  A postoperative protocol was implemented that included IV hydration, nausea control, pain control, incentive spirometry use, vitals assessment, and ability to tolerate PO intake. Patients also had to be assessed by a physician prior to discharge.   - Patients were sent home with a pulse oximeter and spirometer for self-monitoring. All patients were contacted at approximately 10 h postoperatively to assess vital signs and symptoms. - Patients were instructed to call their surgeon directly if they developed a heart rate greater than 100 or a pulse oxygen saturation less than 90%. - Patients returned to the clinic on postoperative day 1 and were seen by a provider, received IV hydration, and labs were obtained. | - 80 of 89 patients (89.8%) were successfully discharged on postoperative day (POD) 0. 3 patients were readmitted within 30 days. - Identification of low-risk patients for outpatient bariatric surgery can be safely accomplished using risk stratification tools. - Implementation of remote monitoring tools can also help support outpatient follow up and improve outcomes. |
| Habib Bedwani, 2023 | Cohort A 191  Cohort B 164 | UK | Prospective, observational | Patients who presented between 26 Mar-6 May 2020 with suspected appendicitis | Not defined | To evaluate the intercollegiate guidelines published in the UK promoting safe surgical care during COVID-19 on appendicitis management.  Patients were identified from a subset of a previous multicentre study as comparator ^87^ . | Within 6 weeks of the new Intercollegiate guidelines,   - 191 patients met the inclusion criteria, of which 79/191 (41%) had appendicitis. - the rate of appendicitis was not statistically different from cohort A 63/164 (38%) - significantly more patients with possible appendicitis underwent CT scanning of the abdomen and pelvis (105/191 vs 71/164) and thorax (66/191 vs 9/164) - there was a significant decrease in ultrasound examinations (62/191 vs 71/164) - significantly fewer patients underwent surgery (52/63 vs 28/79) - there was a statistically significant increase in the number of patients managed non-operatively between cohorts (11/63 vs 51/79) - there were no negative appendicectomies following the introduction of the guidelines compared to 10% prior to the guidelines. - Length of stay was significantly shorter in patients undergoing non-operative compared to operative management (1 day vs 3 days) and was shorter for all patients being treated for appendicitis after introduction of the guidelines (2.5 days vs 2 days) - more patients were followed up in surgical clinics (16/164 vs 58/191) although without a difference in follow-up duration. - There was no statistical difference in complications - there was a significant increase in SARS-CoV-2 PCR testing (9/164 vs 71/191) |
| Jiang, 2021 | 202 | China | Prospective cohort | Elective surgeries conducted during February 11^th^ to March 11^th^ 2020 | Non-elective surgeries conducted outside of February 11^th^ to March 11^th^ 2020 | Remodeled pathways to separate patients and medical personnel during COVID-19, including medical staff and patient education, ward disinfection, screening stations, patient/companion management, and perioperative management.   - Medical staff and patient education: updated our clinical knowledge through medical teleconferencing, and we educated potential day surgery center patients and social media followers through our follow-up application - Ward disinfection: redesigning ward access to separate medical staff and patients, providing sufficient personal protective equipment and other materials (caps, protective gowns, surgical masks, goggles, and disinfectant), and developing a daily shift schedule. - Screening stations: Before entering the day surgery center, patients were required to pass through 3 extra screening stations. Patients were checked for symptoms such as: fever, cough, sore throat, and headache and to determine their epidemiological history to rule out a possible risk of the COVID-19 infection. - Patient/companion management: Patient and companion education was conducted via telephone or follow-up application. They were advised to wash their hands frequently with soap or to sanitize their hands using an alcohol-based sanitizer. They were also advised to avoid touching their eyes, nose, and mouth and to avoid crowded places. They were asked to practice respiratory hygiene; when coughing and sneezing, cover their mouth and nose with a tissue or flexed elbow, throw the tissue away immediately, and wash hands afterward. Last, they were asked to try to avoid close contact with anyone who has a fever and cough. - Medical Staff/Perioperative management: Staff temperature was monitored 3x/day, and were required to change their surgical mask every 4 ours at work. Regular telecommunication continued across the hospital with all clustered meetings conducted by video conference. Postoperative follow-ups continually evaluated patients for 1 month. When patients were discharged, traditional telephone follow-up method was used to contact them on postoperative days 2, 7, 14, and 30 to inquire about symptoms of fever (T ≥ 37.3°C), wound pain, nausea, vomiting, or other complications. | - Days 7, 14, and 30 after discharge, no postoperative complications - Days 7, 14, and 30 after discharge, no positive COVID-19 infections reported |
| Joseph, 2020 | 100 | UK | Randomized & retrospective | Elective Transcatheter Aortic Valve Replacement (TAVR) for the treatment of aortic stenosis. | If referred to the TAVR service greater than one year prior to the date of the planned procedure. | Transcatheter Aortic Valve Replacement (TAVR) was relocated to an ancillary, local private hospital not setup to perform these procedures.  Change in pre- and post-procedural work-up and care and referrals were either direct for TAVR or from the urgent surgical aortic valve re- placement (sAVR) waiting list and the patient was triaged based on high-risk features.  All TAVR procedures were either the transfemoral at the new site or transfemoral/transaxillary access route at the surgical center. The heart valve choice was at the surgeon/operator’s discretion between those available at the center. | No statistically relevant difference between the two treatment periods in the incidence of death or disabling stroke, but there were numerically more minor strokes during the COVID-19 period.  No significant difference in the distribution of procedure length during the COVID-19 pandemic (60 vs 60 min).  There was a significant increase in the proportion of Sapien 3 valves inserted (34 vs 68%).  There was a significant reduction in the length of inpatient stay (3 vs 2 days). |
| Leung, 2021 | 585 | UK | Consecutive (historic) cohorts | Patients undergoing gynecological cancer surgery | N/A | A formal mitigation plan with all relevant steps to reduce COVID-19 infection during open and laparoscopic surgery, along with the surgical modifications introduced.  Steps were formulated at a multidisciplinary consensus meeting with all relevant stakeholders with the organization.  Between February 2020 to April 2020, introduced the use of additional personal protection for our surgical team, and surgical staff was not involved in managing patients with COVID-19 patients. Segregation of wards and operating theatre into COVID-19-free areas.  Cancer service was relocated to a “private sector hospital” (a privately funded hospital that was contracted to undertake NHS work during the COVID-19 pandemic) with capacity to offer post-operative high-dependency care. Importantly, the hospital did not have an emergency department and did not admit or treat patients with suspected or proven COVID-19. Patients were required to self-isolate 14 days prior to surgery and undertake a robust pre-operative health care questionnaire specific for COVID-19 symptoms. From 22^nd^ April 2020, mandatory COVID-19 screening test using  PCR was introduced to all pre-operative patients 3 days prior to their surgery. Surgical team members were fitted with FFP3 masks and eye shields (visors), and surgery was modified to reduce the potential of aerosol transmission during laparotomy or laparoscopic surgery. | - 30-day post-operative complication rates were significantly higher in 2020 than in 2019 (58 [20.1%] versus 32 [10.8%]; p = 0.002). - There were two early post-operative death (≤30 days) in 2020 (versus none in 2019). - Two positive COVID-19 tests during the study period |
| Logishetty, 2021 | 1,142 | UK | Prospective cohort | All adult patients who were on or added to the planned surgery waiting list prior to 28 August 2020, who had not had their operations prior to the cessation of routine surgery. | - Operation was classed as level 1 urgency (required within 24 hours or sooner) - If they were under 17 years of age - If their patient record had incomplete or anomalous data. | Implementation of Surgical Prioritization and Allocation Guidance (SPAG) during COVID-19.  Considered three facets of surgery: safety, biopsychosocial factors, procedural urgency. | - 21 Arm 3 and Arm 4 patients expedited to “Urgent” - No deaths at 30 days post operation. - No admissions to HDU/ICU. - No reported positive COVID-19 tests. |
| Monroy-Iglesias, 2021 |  | Italy/UK |  | All patients undergoing scheduled radical surgery with curative intent for gynaecological, head and neck (H&N), thoracic and urological cancers between 1 March and 30 September 2020  Same cohort in comparable period of 1st March to 30th September 2019 | Outside study period  Only patients with complete data | At IEO, the anaesthetic protocol was devised to minimise aerosol generation and potential exposure to undetected COVID-19 infection in patients with false negative swab tests.   - All the involved staff were required to wear full personal protection equipment (PPE) and only the anaesthetist and nurse had access to the operating theatre during the patient’s anaesthetic procedure. - During the surgery, all staff involved had to remain wearing full PPE throughout the surgical procedure. - Postoperatively, patients were in single rooms with surgical masks and all visiting healthcare professionals were required to use full PPE when entering the room. - Updates on the state and outcome of the patients to family members was given via telephone.   At SELCA, a multidisciplinary team assessed patients’ risk profiles according to new government guidance in relation to their co-morbidities and the potential negative effects of COVID-19.   - If the health risks were deemed too high, patient care was directed to an alternative non-surgical pathway. - The need for a post-operative critical care unit (CCU) bed was evaluated, and if deemed too high and prolonged, alternative treatments were considered. - An enhanced consenting process was utilised, which included agreed levels of care in the postoperative period with some patients electing not to have CCU care if their condition deteriorated after surgery. - Similar to the IEO pathway, all patients were instructed to self-isolate for 14-days to minimise the risk of acquiring COVID-19 infection in the pre-operative period. - Preoperatively, all patients were intubated in the operating theatre with the anaesthetic team wearing full PPE. Once the endotracheal tube was placed, the surgical team waited 20 min before entering the theatre, this was to allow for adequate air exchanges to occur and minimise the exposure to aerosol. - Throughout the surgery, the team was made up of only consultant surgeons, as junior doctors were deployed to other COVID-19 related duties for the first 2 months of the pandemic. Full PPE was adopted by all theatre staff. Another 20 min were taken after the patient was extubated upon completion of the surgical procedure prior to transfer to the recovery room. - There was a mandatory simulation training programme for all theatre staff, which included putting on and removing PPE techniques, intubation techniques and failed intubation drills. | At European Institute of Oncology, IRCCS (IEO)   - There were 1477 radical surgeries with curative intent performed from March to September 2020 (270 for gynaecological, 339 for H&N, 377 for thoracic, and 491 for urological cancers), compared to 1560 surgeries in the same period in 2019 (274 for gynaecological, 350 for H&N, 460 for thoracic, and 476 urological cancers). - There was a decline of 6% in 2020 compared to 2019. The main decline was seen for thoracic surgery where 18% less surgeries were performed in 2020 compared to 2019. - There was a 3% (*n* = 490 vs. 476) increase in urological cancer surgeries in 2020 compared to 2019. - Nine (1%) patients developed COVID-19 post-operatively (1 (<1%) of gynaecological, 7 (2%) of H&N, and 1(<1%) of urological cancer); of these, only the gynaecological patient went on to develop severe disease and died from COVID-19.   At South East London Cancer Alliance (SELCA)   - There was a decline of 34% radical surgeries performed from 23 March to 8 September 2020, compared to the same period in 2019. - There were 1553 radical surgeries in the observed period (321 of breast, 129 of colorectal, 114 of gynaecological, 152 of H&N, 92 of liver, 56 of plastics, 305 thoracic, 72 of upper gastrointestinal (GI), and 312 of urological cancers), compared to 2336 in 2019. - The most notable declines were seen for plastic/skin surgeries, with a decline of 80% (*n* = 56 vs. 278); followed by colorectal with 59% (*n* = 129 vs. 310), and breast with 38% (*n* = 321 vs. 519). - However, there was an increase in the number of H&N (9%, *n* = 152 vs. 139, *p* = 0.00) and upper GI (18%, *n* = 72 vs. 61, *p* = 0.00) cancer surgeries in 2020 compared to 2019.   The current study has shown that the implemented COVID-19 minimal pathways are safe for cancer patients that require radical treatment.  Comparing hospitals with different geographical settings, patient characteristics were comparable between both cancer hubs except for performance status, where in the SELCA population, more surgeries were performed in patients with a lower performance status (0 and 1). |
| Moschovas, 2021 | 147 | USA | Prospective cohort | Oncological treatment, patients undergoing robot-assisted radical prostatectomy | Non-surgical patients undergoing robot- assisted radical prostatectomy (RARP) | Changing the office routine, stratifying the patients according to the National Comprehensive Cancer Network (NCCN) risk, and adopting COVID-19-based criteria to select patients for surgery (based on risk scoring) was an effective means to ensuring quality treatment and minimizing infection.   - Office Routine: Adoption of contact precautions. All staff members were divided in three different shifts to reduce the number of individuals working in the same period. The staff were also required to use masks and gloves during the entire working period, as were the patients during their consultations. All physical contact and object sharing, such as charts and computers in the common areas, were minimized. All patients and employees had their temperature checked and sanitized their hands. - Selection Criteria to surgery: After the division into NCCN groups, the patients were categorized according to a COVID risk stratification based on age, BMI, smoking history, and comorbidities. Patients in the COVID high-risk group had their surgery postponed due to increased mortality and complication rates. - Management of surgical patients: COVID screening is based on questionnaires about the patient’s routine and exposure in the last 14 days and COVID rapid test 48 hours before surgery. Patients were advised to adopt social distancing and quarantine 2 weeks before surgery. Upon arrival on the day of surgery, the patient is given a surgical mask that will remain in place until the surgical intubation. | No intraoperative complications and no postoperative COVID-19 infections |
| Narang, 2020 | 17 | USA | Retrospective chart review | Pregnant patients with anomalous fetuses who met criteria for fetal intervention | Patients who were evaluated but did not undergo an intervention | Beginning March 15, 2020, universal testing for patients admitted to Labor and Delivery was implemented. Screening for those undergoing planned surgical interventions began on April 1, 2020.  The Fetal Surgery During SARS-CoV-2 Pandemic Protocol, adopted from institutional guidelines, including Mayo Clinic perinatal committee guidelines and surgical requirements outlined by the Infection Prevention and Control (IPAC) committee. | This study found no differences between 2019 and 2020 in the number of procedures performed, maternal age, gestational age at procedure, type of anesthesia, intraoperative preparation time, and total procedure time.  There were more fetoscopic laser ablations of the placental anastomoses for severe TTTS during COVID-19 than pre-COVID-19 (66.6% of cases). |
| Ong, 2023 | 554 | USA | Prospective cohort | - Adults aged ≥18 years old - Undergone elective unilateral total knee arthroplasty (TKA) for primary osteoarthritis - Discharged between June 1, 2020, and August 31, 2020 | - Patients who underwent TKA for other diagnoses - TKA with concomitant removal of hardware - Patients who underwent revision procedures | Implementing a modified protocol to evaluate the outcomes and safety of resumption of elective procedures during COVID-19.  Emphasis was placed on shortening hospitalization, discharging patients to their home and increasing the use of telemedicine were techniques utilized in this study. | Using the new protocol, patients:   - Were discharged over one day (25 hours) earlier on average compared to controls - Were discharged earlier, on postoperative day 0 and 1 compared to controls (24.9% vs. 16.1%) - Experienced greater rates of unscheduled outpatient visits (9.2% [51 of 554] vs. 4.9% [27 of 554]) compared to controls, - More frequently used telehealth to attend visits (11.7% [65 of 554] vs. 0% [0 of 554]) - A majority of unscheduled outpatient visits were related to wound complications (51% [26 of 51] and 40.7% [11 of 27]) and postoperative pain/swelling concerns (31.4% [16 of 51] and 44.4% [12 of 27]) - Rates of ER visits (4.5% [25 of 554] vs. 3.2% [18 of 554]) and hospital readmissions (2.9% [16 of 554] vs. 2% [11 of 554]) were similar between cohorts and not significant - Primary motivations for hospital admissions included wound or deep infection (37.5% [6 of 25] and 9.1% [1 of 11]), mechanical (12.5% [2 of 16] and 36.4% [4 of 11]) and pain (25% [4 of 16] and 0% [0 of 11]) related concerns. |
| Pai, 2020 | 593 | USA | Prospective cohort/quality improvement | All pre-operative patients | N/A | A multidisciplinary quality improvement team was formed.  The existing preoperative evaluation (POE) clinic was utilized as a centralized method to provide COVID-19 testing, symptom screening, and infection prevention education in addition to routine preoperative medical optimization.  -During POE triaging calling, nurses provided education on isolation, personal hygiene, and face mask use.  -Patients with symptoms but testing negative for COVID-19 were referred to a COVID-19 Virtual Clinic (CVC) for further evaluation  -If patients tested positive for COVID-19, they were referred to the CVC for telephone or video follow-up and their surgeries were postponed.  DMAIC (define, measure, analyze, improve, control) methodology was used and the study was limited to analyzing scheduling and administrative data. | With the new process, the percentage of patients with COVID-19 testing results returned before surgery increased from 10% to 100%.  Emergent surgeries in which physicians felt a delay in waiting for COVID-19 test results would adversely affect their well-being, would proceed as if they had tested positive.  Of the 593, only 2 patients were found positive during preoperative testing and had their surgeries rescheduled. |
| Ren, 2021 | 4,720 | China | Implementation | Surgical operations | Not defined | Establishment of a graded prevention and control guidance for surgery.  Set up of a dedicated team for prevention and control of COVID-19, and establishment of an emergency system for surgical risk management grounded in risk management theory.  All patients entering the hospital take a temperature test. Before entering other departments (outpatient, emergency) the precheck triaging team will also conduct temperature test and ask the patient’s history.  The risk assessment focuses on two opportunities:  1. The preoperative evaluation of the doctor in charge  2. The assessment of the anesthesiologist and the operating nurse  According to the results of the evaluation, different levels of prevention and control measures should be implemented accurately during the operation.  Postoperative management included:   - Personnel management - Environmental disinfection - Changing the high efficiency particulate air filter - Processing of reusable surgical instruments - Health care waste treatment | The surgical prevention measures-based risk stratification included prescreening and preoperative risk assessment, preparation of operating room, medical staff protection and environmental disinfection measures.  There were 1,565 emergency operations and 22 for medium-risk and high-risk patients, without confirmation of COVID-19 testing.  There were no medical staff exposed during the implementation of protective measures. |
| Sebastian, 2022 | 764 | UK | Cohort | Adults aged ≥18 years old  Acute severe ulcerative colitis (ASUC) fulfilling Truelove and Witts criteria  ASUC diagnosis between either 1 March 2020 and 30 June 2020 (COVID-19 pandemic period) or 1 January 2019 and 30 June 2019 (prepandemic period). | Patients who did not meet inclusion criteria  Patients with Crohn’s disease, cytomegalovirus or Clostridium difficile infections | Instead of patients being admitted for treatment of acute severe ulcerative colitis (ASUC) and treated with IV therapy and/or colectomy, an alternative form of treatment was administered that did not involve the above.  Patients were administered IV corticosteroids or rescue therapy in the ambulatory setting. | Patients managed as ambulatory from diagnosis received therapy sooner than the those who were inpatient or discharged to ambulatory pathways.  The response to induction therapy is notably lower in patients managed as ambulatory from primary diagnosis.  Patients discharged to ambulatory pathways were most likely to undergo a colectomy sooner.  The mortality rate is greatest in the largest cohort, those receiving inpatient care.  The length of stay for those patients managed as ambulatory from diagnosis is greatest.  Those receiving inpatient care were most likely to undergo a colectomy within the 3-mo f/up period. |
| Sivaraj, 2022 | 711 | UK | Prospective cohort | Minimally invasive surgery, open surgery, and endoscopic surgeries | Non-surgical procedure patients | Observing the safety of minimally invasive surgeries compared to open surgeries, and comparing safety between COVID-19-10 lockdowns and pre-COVID-19 2019. | 8.5% of MIS 2020 had respiratory complications (2019: 7.7%, 2021: 6.9%; p = 0.9) vs 10.5% in OS 2020 (p = 0.8 vs MIS)  Median LOS[IQR] for MIS 2020 was 2.5[6] days vs 5[23] days in OS 2020 (p = 0.06) |
| Stagg, 2022 | 72 | USA | Prospective cohort | Infants with   - shunt-dependent single ventricle physiology (surgical shunt placement or PDA stenting as part of staged palliation) - single ventricular physiology and a pulmonary artery band - aortopulmonary shunts - PDA stents   in anticipation of eventual biventricular repair  AND  infants with stable neonatal physiology with either:   - single ventricle physiology - those with undetermined candidacy for biventricular repair with the above interventions | Infants without   - shunt-dependent single ventricle physiology (surgical shunt placement or PDA stenting as part of staged palliation) - single ventricular physiology and a pulmonary artery band - aortopulmonary shunts - PDA stents   OR  infants without stable neonatal physiology with neither:   - single ventricle physiology - those with undetermined candidacy for biventricular repair with the above interventions | Replaced at least one in-person primary care provider visit with a telemedicine cardiology visit.  The telemedicine includes a pediatric cardiologist, nurse practitioner, and patient’s primary cardiologist including fellows. | Significantly lower number of ED visits compared to the same calendar period of the prior year (0.0 (0–2.5) vs. 0.4 (0–3.7), p = 0.0004) |
| Turkdogan, 2023 | 17 | Canada | Prospective | All percutaneous tracheotomies (PT) in the ICU between the months of April 2020 – January 2021 | Tracheostomies performed outside the study period or outside the ICU | Creation of a novel method for a safe surgical environment to conduct a tracheostomy. This method covered the surgical field with a modified demistifier canopy.  A demistifier canopy is a large, clear canopy often used in the treatment of patients with respiratory syncytial virus (RSV) infection. It is readily available, inexpensive, and simple to modify and install.  The method created a specialized tracheostomy team was mobilized, consisting of a surgeon, a critical care physician, 2 respiratory therapists, 3 senior residents, and experienced ICU nurses.  Before conducting the intervention, a simulation was performed two-days prior. Every step was practiced multiple times, and every individual’s role was defined. | There were no procedure-related complications, and no evidence of COVID-19 transmission to any member of the health care team during the study period.  The study believe the modified PT technique provides superior safety and allows easier and timelier interventions in intubated COVID-19 positive patients. |
| Vaidya, 2021 | N/A | USA | Focus group/expert consensus  Implementation process | Inclusion in the Connecticut Orthopaedic Institute | N/A | Pivot Plan revolved around four domains: safety, space, staff, and supplies.  The plan was implemented across three phases of surgical care:  Phase I: preoperative care   - Virtual preoperative education was initiated. A virtual online class was started at MidState Medical Center (MMC), while St. Vincent’s Medical Center (SVMC) began to offer a navigator-led class via telephone. As part of the virtual education, patients gained online access to educational materials available anytime from the convenience of their homes. MMC also began to offer live virtual webinars in addition to the existing online preoperative classes. - A clinical command center was also established to facilitate scheduling of preoperative testing. In addition, a system-wide COVID-19 “dashboard” was developed in the electronic health record (EHR) to easily access the results of COVID-19 testing. - The preadmission testing center (PAC) was established as the initial patient contact after the patient was scheduled for surgery. The primary goals were to 1) risk-stratify patients preoperatively and facilitate consults as indicated, 2) call patients one to two weeks before surgery to facilitate medical clearance by offering history and physical (H&P) and preadmission testing, 3) address any last-minute test-related questions or concerns and reinforce hospital safety measures, 4) offer COVID-19 polymerase chain reaction (PCR) testing within seven days of the date of surgery, 5) ensure patients remain in self-quarantine in-between the COVID-19 test and the procedure date, and 6) alert patients to any potential physical findings on the day of surgery which may lead to postponement or cancellation of surgery. - Upon resumption of elective orthopaedic surgeries, orthopaedic navigators at MMC and SVMC performed risk assessment evaluations according to system-wide criteria. - Marketing began in June 2020, with 1) mailed flyers containing hospital safety data to patients who had not rescheduled elective surgeries, 2) followed up with patients via patient navigators, 3) posted recent patient testimonials about their experiences during the pandemic on the COI website, 4) created a 30-second advertisement addressing COVID-19 safety and the COI’s commitment to delivering a high level of care to our patients, 5) went live with providers on social media platforms and gave interviews on a local news station to reiterate COVID-19 safety, and 6) developed a safety message video for social media highlighting some of the COI amenities, as well as COVID-19 precautions.   Phase II: inpatient care   - Patients were required to wear masks to the hospital and began to be temperature and symptom screened at the front entrance (e.g., fever, cough, shortness of breath, gastrointestinal upset, etc.). - All staff was required to wear an N95 respirator in the operating room. This requirement changed in July 2020 and was no longer required for patients with a documented negative COVID-19 test. - Staggered patient bays in the post-anesthesia care unit (PACU) were used if sufficient room was available, and if bays held two patients, the curtain was utilized to separate the patients. Six-foot social distancing was implemented throughout the care process. Same-day surgical patients recovered and were transitioned to home from recovery spaces - A system-wide checklist for safety was created, scaling, and sustaining best practices for infection prevention. Checklist elements included PPE and infection prevention, facilities/infrastructure, team member management, patient monitoring and screening, visitor monitoring and screening, performance management, and quality control.   Phase III: postoperative discharge care   - Discharge transportation was arranged to observe social distancing rules. Post- discharge instructions regarding COVID-19 began to populate the discharge summary automatically. - Increased touch-points between the navigator and patients were used to mitigate any complications, questions, or concerns that may occur postoperatively. - Discharge planning staff began to receive weekly updates on the operation and percentage of COVID-19-positive patients at all local skilled nursing facilities. | Caseloads and OR turnaround time were comparable to those pre-COVID-19 with implementation of the Pivot Plan |
| Wee, 2020 | 8,437 | US | Cohort | Surgical patients admitted with concurrent respiratory symptoms, infiltrates on chest imaging or suspicious travel/epidemiologic history from January to April 2020 | Patients without clinical signs or symptoms or patients suspected outside the study period | Risk-stratified approach to conduct peri-operative testing for COVID-19. | The testing yield was lower in surgical inpatients compared with medical inpatients.  A risk-stratified testing strategy picked up previously unsuspected COVID-19 in six cases; 66.7% (4/6) were asymptomatic at presentation. |
| Wienhold, 2021 | 111 | Germany | Implementation | Elective surgery | Language or cognitive barriers  High-risk procedures with need for postoperative ICU stay  Acute infection with required auscultation | Development of a teleconsultation preoperative evaluation platform | 93.7% of anaesthetists rated the telepreoperative evaluations as equivalent to on-site meetings.  Most patients considered teleconsultation for preoperative evaluation as convenient as an on-site meeting (98.2%) and would choose a teleconsultation again (97.9%).  Median travel time saved by patients was 60min (Q1 40, Q3 80).  There was one adverse event detected: atrial fibrillation only immediately prior to surgery. |
| Yap, 2023 | 77 | Singapore | Prospective comparator | - age ≥5 years - abdominal pain duration ≤48 h - ultrasound (US)/Computer Tomography scan confirmation of AUA   US appendiceal diameter 6–11 mm with no features of perforation/abscess collection and no faecolith | - patients with generalized peritonitis on clinical examination   patients with a significant medical history or concurrent medical condition that may potentially place them at a higher risk for complications | Eligible patients were counselled regarding both treatment options – upfront surgery versus non-operative treatment (NOT) with antibiotics alone.  -For patients managed with NOT, initial intravenous (IV) antibiotics, Amoxicillin/Clavulanic acid or a combination of Ceftriaxone and Metronidazole were administered for a minimum of 24 h. Patients were discharged home with a course of oral antibiotics for 10 days. On discharge, patients were given a diary to record their home recovery.  For patients managed through laparoscopic appendectomy, received IV antibiotics (either Amoxicillin/Clavulanic acid or Ceftriaxone and Metronidazole) for 24 hours perioperatively. Patients were discharged when able to resume normal diet and were without early post-operative complications. | Success of NOT at index admission was 90.7% (39/43).  NOT failure rate at 27 months’ follow-up was 37.2% (16/43).  Of the NOT failures, 1 appendix was normal on histology while only 1 was perforated.  There were no significant differences in secondary outcomes between both groups except for LOS of late NOT failure. |
| Zago, 2020 | 49 | Italy | Non-experimental | Patients undergoing surgery | N/A | Two systems including the same type of filter were developed:   1. Use of a male-to-male connector, a 25-cm long Luerlock infusion connector, an electrostatic filter HME (Heat and Moisture Exchanger) with end tidal CO2 port, and a 15-mm cap for fenestrated inner tracheostomy cannula 2. A rubber/Luer-lock connector, commonly used for infusion of fluids or connecting drainages; a blue connector included in the Rüsch™ Breathing kit for ventilators, and the same electrostatic filter HME.   The key object of these constructions is the filter. HME filters have high resistance to flow and, most important, a bacterial and viral filtration efficiency of ≥ 99.999%. | Both systems are believed to be effective in surgical populations, and not surgical personnel have tested positive for COVID-19 |

#### Pressure Injury

| **Au, Yr** | **Patients, N** | **Country** | **Study design** | **Inclusion criteria** | **Exclusion criteria** | **Description of intervention, innovation, adaptation, or change in practice** | **Narrative Outcome Description** |
| --- | --- | --- | --- | --- | --- | --- | --- |
| Doussot, 2020 | 117 | France | Prospective cohort | Patients admitted to ICU requiring prone positioning | Patients not requiring prone positioning | After accelerated simulation training, 109 volunteers including surgeons, physicians, nurses and physiotherapists, multiple dedicated teams performed daily multiple prone positioning (PP) following a systematic checklist. | Of the 117 patients,  67 patients (57.3%) required prone positioning.  A total of 384 PP were performed. Overall, complication occurred in 34 prone positioning (8.8%) and led to PP cancelation in 4 patients (1%).  Four health care workers presented with potential COVID-19 related symptoms and none was positive. |
| Gallagher, 2022 | 100 or more | US | Cohort | Patients with COVID-19 required protracted  mechanical ventilation necessitating tracheostomy | Not stated, but would exclude those without tracheostomy | Implementation of hospital-wide bedside rounds on all adults with tracheostomies. Skin and safety assessments were performed with peer-to-peer coaching. Data | Upon initiation of rounds in June 2020:   - Only 49% of patients had an obturator at the bedside - 82% of patients had a backup tracheostomy at the bedside - 72% of patients had a preventive dressing in place.   By December 2020, safety measures had improved:   - 97% of patients had an obturator at the bedside - 90% of patients had a backup tracheostomy at the bedside - 96% of patients had a preventive foam dressing in place.   At the time of the COVID-19 surge:   - Pressure injury from tracheostomy was 9, 19, and 4 case in April, May and June. - After the intervention, rate of pressure injury has ranged from 0-4 cases over the following 6 months |
| Miguel, 2021 | 147 patients turned 450 times | US | Descriptive | N/A | N/A | Creation and training of proning team (PT) .  Using the procedure developed by this institution’s ICU clinical nurse specialists, a new curriculum titled “Proning Intubated Patients in the Intensive Care Unit” was developed.  Training included:   - A review of the purpose, indications, expected outcomes, and potential complications of PP, a step-by-step video of the procedure, and a refresher - on proper technique for donning and doffing personal protective equipment. - A 60-minute simulation, led by the critical care clinical nurse specialist, allowed trainees to practice the basic safety steps for manually turning a patient receiving ventilatory support to prone position and back to supine position.   The PT was available 24/7. There were 2 teams of 4 on both the day and evening shifts, and 1 team of 3 on the night shift.  Each PT consisted of a minimum of 1 operating room registered nurse and 1 physical therapist.  The PT’s supportive tools included:   - daily report sheets for rounding to all ICUs; - real-time reports from the electronic medical record identifying all patients with COVID-19 who were receiving ventilatory support and their basic demographics (ie, age, sex, and weight); - an enhanced checklist for preproning, proning, and postproning needs. | The PT successfully turned 147 patients 450 times without loss of any oral or endotracheal airways; arterial, venous, or central catheters; or any tubes and drains.  No team member sustained physical injuries or COVID-19–positive conversions. |
| Rana, 2021 | 524 patients | USA | Implementation | Patients admitted to ICU | Patients < 18 years of age  Patients admitted after an elective procedure or surgery  Patients with COVID-19 | Fall “visits” were completed with direct care providers for the team to examine the variations between work-as-  imagined (what was expected to happen) and work-as-done (what actually happened) to improve uptake and ease of retention by frontline  staff. | The average inpatient fall rate in the hospital of this study was 0.630 per 1000 patient bed days from January 2019 to June 2020. |
| Taylor, 2021 | 40 | USA | Clustered, randomized pilot trial | Patients admitted to the hospital by one of the study teams, had tested positive for SARS-CoV-2 within 7 days or were suspected to have COVID-19 pneumonia, and experienced  1) room air  oxygen saturation,93% or  2) oxygen requirement of 3 L per minute or greater  without the need for mechanical ventilation | Patients unable to self-turn, spinal  instability, facial or pelvic fractures, open chest  or abdomen, altered mental status, anticipated  difficult airway, signs of respiratory fatigue, or  receiving end-of-life care) | - Delivery of prone positioning education and explanation of risks and benefits to patients by bedside clinicians. - Routine monitoring for worsening status; - Attempts to improve comfort as needed. - Patients were encouraged to sustain the prone position as long as possible but were allowed to return to the supine position as necessary | None of the patients could remain prone for the12-to 16-h prone target time suggested by clinicians with patients estimated target time suggested by clinicians with patients estimated spending between 10 and 120 min a day in prone position.  Our results indicate that assessing fidelity to prone positioning is a major challenge in a pragmatic study design using data captured for clinical use. |

#### Falls

| **Au, Yr** | **Patients, N** | **Country** | **Study design** | **Inclusion criteria** | **Exclusion criteria** | **Description of intervention, innovation, adaptation, or change in practice** | **Narrative Outcome Description** |
| --- | --- | --- | --- | --- | --- | --- | --- |
| Kwok, 2022 | 347  falls | China | Retrospective | Inpatient falls | Not defined | Formation of a hospital falls review team composed of a geriatrician, nurses, physiotherapists, occupational therapist, human factors specialist (a hospital patient safety officer chartered in HF&E) and administrators.  Creation of a falls prevention programme was developed in July 2020.  Upon notification of a fall, the Team conducted a short visit on the ward on the incident day or next working day.  The elements on each category of the framework was used by the visiting members as quick prompts of areas to inspect. The visiting members reviewed and discussed with ward staff about the fall preventive measures applied on the patient.  The visit reviewed the patient records related to falls prevention and postfall management.  The team also conducted special visits to inpatient wards (both in service and under renovation) to assess the equipment, environmental and workflow design on falls prevention when requested by clinical departments.  Fall summaries were created that summarized the discussion of observations of each fall for monthly meetings with a multidisciplinary approach. | A total of 120, 85 and 142 inpatient falls in the ‘pre-COVID-19’, ‘COVID-19’ and ‘programme’ periods were reviewed, respectively. Thirteen areas of observations were identified by the Team.  When reviewing the falls, the team adopted a systems approach such that most actions taken were believed to be generalisable to other departments or wards in the hospital.  The average fall rates were found to be significantly increased from the ‘pre-COVID-19’ to ‘COVID-19’ periods (mean difference=0.297), while a significant decrease was noted between the ‘COVID-19’ and ‘programme’ periods (mean difference=−0.226). |
| Rana, 2021 | 524 | USA | Implementation | Patients admitted to ICU | Patients < 18 years of age  Patients admitted after an elective procedure or surgery  Patients with COVID-19 | Fall “visits” were completed with direct care providers for the team to examine the variations between work-as-  imagined (what was expected to happen) and work-as-done (what actually happened) to improve uptake and ease of retention by frontline  staff. | The average inpatient fall rate in the hospital of this study was 0.630 per 1000 patient bed days from January 2019 to June 2020. |
