## Supplementary material for "INTERVENTIONS TO IMPROVE PATIENT SAFETY DURING THE COVID-19 PANDEMIC: A SYSTEMATIC REVIEW": Annex B

### Annex B: Data Abstraction Tables

#### Annex B1. Skeleton for evidence tables

| Au, yr | Patients, n | Country | Study Design | Inclusion criteria | Exclusion criteria | Description of intervention, innovation, adaptation, or change in practice | Narrative Outcome Description |
| --- | --- | --- | --- | --- | --- | --- | --- |
