## Supplementary material for "INTERVENTIONS TO IMPROVE PATIENT SAFETY DURING THE COVID-19 PANDEMIC: A SYSTEMATIC REVIEW": Annex A

### Annex A: Detailed PubMed Search Strategies

**A1: PubMed Search strategy for medication safety.**

| Search # | String |
| --- | --- |
| 1 | "Patient Safety"[Mesh] OR "patient safety"[tiab] OR "patient safety"[mh] OR "risk management"[tiab] OR "risk management"[mh] OR “safety incident”[tiab] OR “safety incidents”[tiab] |
| 2 | "Medication Errors"[Mesh] OR "medication error"[tiab] OR "medication errors"[tiab] OR ("Look Alike Sound Alike"[tiab] AND error[tiab]) OR (Prescriptions[mh] AND (error[tiab] OR errors[tiab])) OR “prescription error”[tiab] OR “prescription errors”[tiab] OR “drug error”[tiab] OR “drug errors”[tiab] OR “Inappropriate Prescribing”[mh] OR “inappropriate prescribe*”[tiab] OR “dose failure”[tiab] OR “dosing error”[tiab] OR “dosing errors”[tiab] OR “dose error”[tiab] OR “dose errors”[tiab] OR (“inappropriate selection”[tiab] AND (drug[tiab] OR medication[tiab])) OR “Drug interaction”[tiab] OR “Drug interactions”[tiab] |
| 3 | "COVID-19"[Mesh] OR "COVID-19"[tiab] OR "SARS-CoV-2"[tiab] OR "2019 Novel Coronavirus"[tiab] OR "Coronavirus Disease 2019"[tiab] OR "Pandemics"[Mesh] OR pandemics[tiab] OR pandemic[tiab] |
| 4 | "Case Reports" [Publication Type] OR "Editorial" [Publication Type] OR "Letter" [Publication Type] OR "Comment" [Publication Type] OR "Organizational Case Studies"[Mesh] OR “case report”[tiab] OR “case reports”[tiab] OR “case study”[tiab] OR “case studies”[tiab] OR “case history”[tiab] OR “case histories” OR “case series”[tiab] OR editorial[tiab] OR editorials[tiab] OR “letter to the editor”[tiab] OR comment[tiab] OR commentary[tiab] |
| 5 | (#1 AND #2 AND #3) NOT #4—limited to studies published March 2020 through 5 October 2022 |

**A2: PubMed search strategy for diagnostic safety.**

| Search # | String |
| --- | --- |
| 1 | “Patient Safety”[Mesh] OR “patient safety”[tiab] OR “patient safety”[mh] OR “risk management”[tiab] OR “risk management”[mh] OR “safety incident”[tiab] OR “safety incidents”[tiab] |
| 2 | “Diagnostic Errors”[MH] OR “Delayed Diagnosis”[MH] OR “diagnostic error”[tiab] OR “diagnosis error”[tiab] OR “delayed diagnosis”[tiab] OR “diagnostic errors”[tiab] OR “diagnosis errors”[tiab] OR misdiagnosis[tiab[ OR “missed diagnosis”[tiab] OR misdiagnoses[tiab] OR “missed diagnoses”[tiab] OR “wrong diagnosis”[tiab] OR “wrong diagnoses”[tiab] OR “inaccurate diagnosis”[tiab] OR “inaccurate diagnoses”[tiab] OR “delayed diagnosis”[tiab] OR “delayed diagnoses”[tiab] OR “diagnosis delay”[tiab] OR “diagnosis delays”[tiab] OR “diagnostic delay”[tiab] OR “diagnostic delays”[tiab] OR “failure to diagnose”[tiab] OR “diagnostic interval”[tiab] OR “diagnostic intervals”[tiab] OR (diagnos*[tiab] AND delay*[tiab]) |
| 3 | “Case Reports” [Publication Type] OR “Editorial” [Publication Type] OR “Letter” [Publication Type] OR “Comment” [Publication Type] OR “Organizational Case Studies”[Mesh] OR “case report”[tiab] OR “case reports”[tiab] OR “case study”[tiab] OR “case studies”[tiab] OR “case history”[tiab] OR “case histories” OR “case series”[tiab] OR editorial[tiab] OR editorials[tiab] OR “letter to the editor”[tiab] OR comment[tiab] OR commentary[tiab] |
| 4 | (#1 AND #2) NOT #3—limited to studies published March 2020 through 5 October 2022 |

**A3: PubMed search strategy for surgical safety.**

| Search # | String |
| --- | --- |
| 1 | "Patient Safety"[Mesh] OR "patient safety"[tiab] OR "patient safety"[mh] OR "risk management"[tiab] OR "risk management"[mh] OR “safety incident”[tiab] OR “safety incidents”[tiab] OR delay[tiab] OR delays[tiab] OR delayed[tiab] |
| 2 | ((Surgical[tiab] OR surgery[tiab]) AND (error[tiab] OR errors[tiab] OR safety[tiab] OR mistake[tiab] OR mistakes[tiab] OR "Cross Infection"[MH] OR “cross infection”[tiab] OR complication[tiab] OR complications[tiab] OR delay[tiab] OR delays[tiab] OR delayed[tiab])) OR “surgical site infection”[tiab] |
| 3 | "COVID-19"[Mesh] OR "COVID-19"[tiab] OR "SARS-CoV-2"[tiab] OR "2019 Novel Coronavirus"[tiab] OR "Coronavirus Disease 2019"[tiab] OR "Pandemics"[Mesh] OR pandemics[tiab] OR pandemic[tiab] |
| 4 | "Case Reports" [Publication Type] OR "Editorial" [Publication Type] OR "Letter" [Publication Type] OR "Comment" [Publication Type] OR "Organizational Case Studies"[Mesh] OR “case report”[tiab] OR “case reports”[tiab] OR “case study”[tiab] OR “case studies”[tiab] OR “case history”[tiab] OR “case histories” OR “case series”[tiab] OR editorial[tiab] OR editorials[tiab] OR “letter to the editor”[tiab] OR comment[tiab] OR commentary[tiab] |
| 5 | (#1 AND #2 AND #3) NOT #4—limited to studies published March 2020 through 5 October 2022 |

**A4: PubMed search strategy for health care associated infections.**

| Search # | String |
| --- | --- |
| 1 | “Patient Safety”[Mesh] OR “patient safety”[tiab] OR “patient safety”[mh] OR “risk management”[tiab] OR “risk management”[mh] OR “safety incident”[tiab] OR “safety incidents”[tiab] |
| 2 | “Cross Infection”[Mesh] OR “cross infection”[tiab] OR “cross infections”[tiab] OR “health care associated infection”[tiab] OR “health care associated infection”[tiab] OR “health care associated infections”[tiab] OR “health care associated infections”[tiab] OR “hospital infection”[tiab] OR “hospital infections”[tiab] OR “Nosocomial Infection”[tiab] OR “Nosocomial Infections”[tiab] OR “Catheter-Related Infections”[mh] OR “Clostridioides difficile”[MH] OR “Drug Resistance, Multiple”[mh] OR “Clostridioides difficile”[tiab] OR “central line associated blood stream infection”[tiab] OR “catheter associated urinary Tract infection”[tiab] OR “surgical site infection”[tiab] OR (“drug resistance”[tiab] AND multiple[tiab]) |
| 3 | “Case Reports” [Publication Type] OR “Editorial” [Publication Type] OR “Letter” [Publication Type] OR “Comment” [Publication Type] OR “Organizational Case Studies”[Mesh] OR “case report”[tiab] OR “case reports”[tiab] OR “case study”[tiab] OR “case studies”[tiab] OR “case history”[tiab] OR “case histories” OR “case series”[tiab] OR editorial[tiab] OR editorials[tiab] OR “letter to the editor”[tiab] OR comment[tiab] OR commentary[tiab] |
| 4 | (#1 AND #2) NOT #3—limited to studies published March 2020 through 5 October 2022 |

**A5: PubMed search strategy for pressure injury.**

| Search # | String |
| --- | --- |
| 1 | “Patient Safety”[Mesh] OR “patient safety”[tiab] OR “patient safety”[mh] OR “risk management”[tiab] OR “risk management”[mh] OR “safety incident”[tiab] OR “safety incidents”[tiab] |
| 2 | “Pressure Ulcer”[Mesh] OR “pressure ulcer”[tiab] OR “pressure ulcers”[tiab] OR “pressure injury”[tiab] OR “pressure injury”[tiab] OR bedsore[tiab] OR bedsores[tiab] OR “bed sore”[tiab] OR “bed sores”[tiab]  OR “pressure sore”[tiab] OR “pressure sores”[tiab] OR “Decubitus Ulcer”[tiab] OR “Decubitus Ulcers”[tiab] |
| 3 | “Case Reports” [Publication Type] OR “Editorial” [Publication Type] OR “Letter” [Publication Type] OR “Comment” [Publication Type] OR “Organizational Case Studies”[Mesh] OR “case report”[tiab] OR “case reports”[tiab] OR “case study”[tiab] OR “case studies”[tiab] OR “case history”[tiab] OR “case histories” OR “case series”[tiab] OR editorial[tiab] OR editorials[tiab] OR “letter to the editor”[tiab] OR comment[tiab] OR commentary[tiab] |
| 4 | (#1 AND #2) NOT #3—limited to studies published March 2020 through 5 October 2022 |

**A6: PubMed search strategy for falls.**

| Search # | String |
| --- | --- |
| 1 | “Patient Safety”[Mesh] OR “patient safety”[tiab] OR “patient safety”[mh] OR “risk management”[tiab] OR “risk management”[mh] OR “safety incident”[tiab] OR “safety incidents”[tiab] |
| 2 | "Accidental Falls"[Mesh] OR falls[tiab] OR fall[tiab] OR falling[tiab] OR slip[tiab] |
| 3 | “Case Reports” [Publication Type] OR “Editorial” [Publication Type] OR “Letter” [Publication Type] OR “Comment” [Publication Type] OR “Organizational Case Studies”[Mesh] OR “case report”[tiab] OR “case reports”[tiab] OR “case study”[tiab] OR “case studies”[tiab] OR “case history”[tiab] OR “case histories” OR “case series”[tiab] OR editorial[tiab] OR editorials[tiab] OR “letter to the editor”[tiab] OR comment[tiab] OR commentary[tiab] |
| 4 | (#1 AND #2) NOT #3—limited to studies published March 2020 through 5 October 2022 |
