## Supplementary material for "INTERVENTIONS TO IMPROVE PATIENT SAFETY DURING THE COVID-19 PANDEMIC: A SYSTEMATIC REVIEW": Annex F

### Annex F: Scale for the quality assessment of narrative review articles (SANRA) assessment.

Table 6: Scale for the quality assessment of narrative review articles (SANRA) assessment^12^.

| **Question** | Answer | **Score** |
| --- | --- | --- |
| **1) Justification of the article's importance for the readership** | The importance is not justified: 0  The importance is alluded to, but not explicitly justified: 1  The importance is explicitly justified: 2 | **2** |
| **2) Statement of concrete aims or formulation of questions** | No aims or questions are formulated: 0  Aims are formulated generally but not concretely or in terms of clear questions: 1  One or more concrete aims or questions are formulated: 2 | **2** |
| **3) Description of the literature search** | The search strategy is not presented: 0  The literature search is described briefly: 1  The literature search is described in detail, including search terms and inclusion criteria: 2 | **2** |
| **4) Referencing** | Key statements are not supported by references.: 0  The referencing of key statements is inconsistent: 1  Key statements are supported by references: 2 | **2** |
| **5) Scientific reasoning**  *e.g., absolute vs relative risk; effect sizes without confidence intervals* | Data are presented inadequately: 0  Data are often not presented in the most appropriate way: 1  Relevant outcome data are generally presented appropriately: 2 | **2** |
